# Human Islet Transcriptomics in Type 2 Diabetes Reveals Conserved Dysfunction Axes and a Candidate Gene Panel

**DOI:** 10.64898/2026.06.08.26355184

**Authors:** Ricardo Romero

## Abstract

**Background.:** Type 2 diabetes mellitus (T2D) involves progressive pancreatic beta-cell dysfunction, but conserved islet programs require cross-cohort and single-cell validation.

**Methods.:** We integrated four human islet transcriptomic cohorts using differential expression, random-effects meta-analysis, Hallmark scoring, and module scoring. To test generalizability without reusing held-out outcomes, modules were re-derived from three cohorts and scored in the fourth in rotating study-level cross-fitting, with expression-and detection-matched random gene-set nulls. Machine-learning performance in GSE164416 was estimated by nested stratified 5-fold cross-validation with DEG calling and feature selection repeated within each outer training fold. An annotation-compatible score was tested in GSE50244, and beta-cell validation used GSE221156.

**Results.:** Cross-fitted IsletDysfunctionScore effects were positive in all four held-out cohorts (d = 0.39-1.36; pooled Hedges g = 0.905, 95% CI 0.501-1.309, p = 1.14 x 10^-5, I2 = 5.5%). Matched gene-set null testing supported effects beyond random expectation in two cohorts. Nested machine learning yielded AUC = 0.960 (bootstrap 95% CI 0.900-0.997); GABRA2 and ENSG00000284653 were selected in all outer folds. The external 8-gene consensus-pool score achieved AUC = 0.907 (DeLong 95% CI 0.818-0.996), but its fixed threshold did not transfer. In GSE221156, the single-cell dysfunction-score analog increased and BetaIdentitySecretion decreased in T2D; ImmuneStress was nominally increased but did not survive false-discovery correction across the single-cell score family.

**Conclusions.:** Human T2D islets show a reproducible stress/identity-loss framework that generalizes directionally under study-level cross-fitting and beta-cell-resolved validation. The candidate panel is a research transcriptomic signature requiring further validation.

## 1. INTRODUCTION

Type 2 diabetes mellitus (T2D) is a chronic metabolic disorder defined by progressive pancreatic beta-cell failure in the context of insulin resistance, affecting hundreds of millions of adults globally and projected to increase substantially over the coming decades (1). Although clinical diagnosis relies on glycaemic thresholds, beta-cell dysfunction develops before overt diagnostic classification and reflects coordinated changes in cellular identity, secretory competence, inflammatory stress, and metabolic resilience (2–5).

A central insight from human islet studies is that T2D is not simply a state of reduced insulin output. Beta cells can lose mature identity features and adopt dysfunctional or partially dedifferentiated transcriptional programs (3). This matters because transcriptional reprogramming may be partially reversible, whereas irreversible loss of beta-cell mass is harder to treat. At the same time, islet inflammation, oxidative stress, donor metabolic state, and tissue-handling factors complicate interpretation of bulk islet transcriptomes.

Public human islet resources have expanded substantially through programs such as the Human Pancreas Analysis Program (HPAP), HPAP-T2D, the Integrated Islet Distribution Program (IIDP), and related deeply phenotyped donor cohorts (6–9). Bulk and microarray datasets remain valuable for cross-platform reproducibility because they capture independent laboratories, platforms, and donor collections. However, bulk isolated-islet profiles cannot fully separate beta-cell-intrinsic transcriptional remodeling from shifts in cell composition, exocrine carryover, vascular/stromal components, or immune infiltration. Laser-capture microdissection improves endocrine enrichment but should not be equated with purified single-cell beta-cell transcriptomes.

Single-cell and single-nucleus RNA-seq now allow cell-type-resolved validation of bulk-derived signatures. Large islet single-cell atlases provide a direct way to ask whether a bulk islet dysfunction program is present within beta cells themselves. In particular, GSE221156 provides a 10x single-cell atlas of 245,878 islet cells from 48 donors spanning non-diabetic, prediabetic, and T2D states, including a reintegrated beta-cell dataset of 99,029 beta cells (9–10). This dataset directly addresses whether the identity-loss component of an islet signature is beta-cell intrinsic. Machine learning (ML) applied to transcriptomic data can produce compact candidate panels, but near-perfect internal performance in high-dimensional omics does not establish universality. Independent validation must assess not only discrimination but also calibration and threshold transfer, because a score may preserve rank-based separation while failing to retain an operating cutoff across platforms or donor collections (11–14). For this reason, this study uses conservative terminology: the 10-gene model is a candidate transcriptomic panel, not a clinically validated diagnostic test.

Here, we combine four complementary layers: (i) cross-platform meta-analysis of public human islet cohorts, (ii) compact immune/stress and beta-cell identity/secretion modules with study-level cross-fitted validation, (iii) nested machine-learning feature selection and external transportability analysis, and (iv) independent single-cell beta-cell validation. We hypothesized that a conserved islet dysfunction program would retain its direction in cohorts excluded from module definition and that its identity-loss component would be recoverable within beta cells at single-cell resolution (Figure 1).

**Figure 1.**
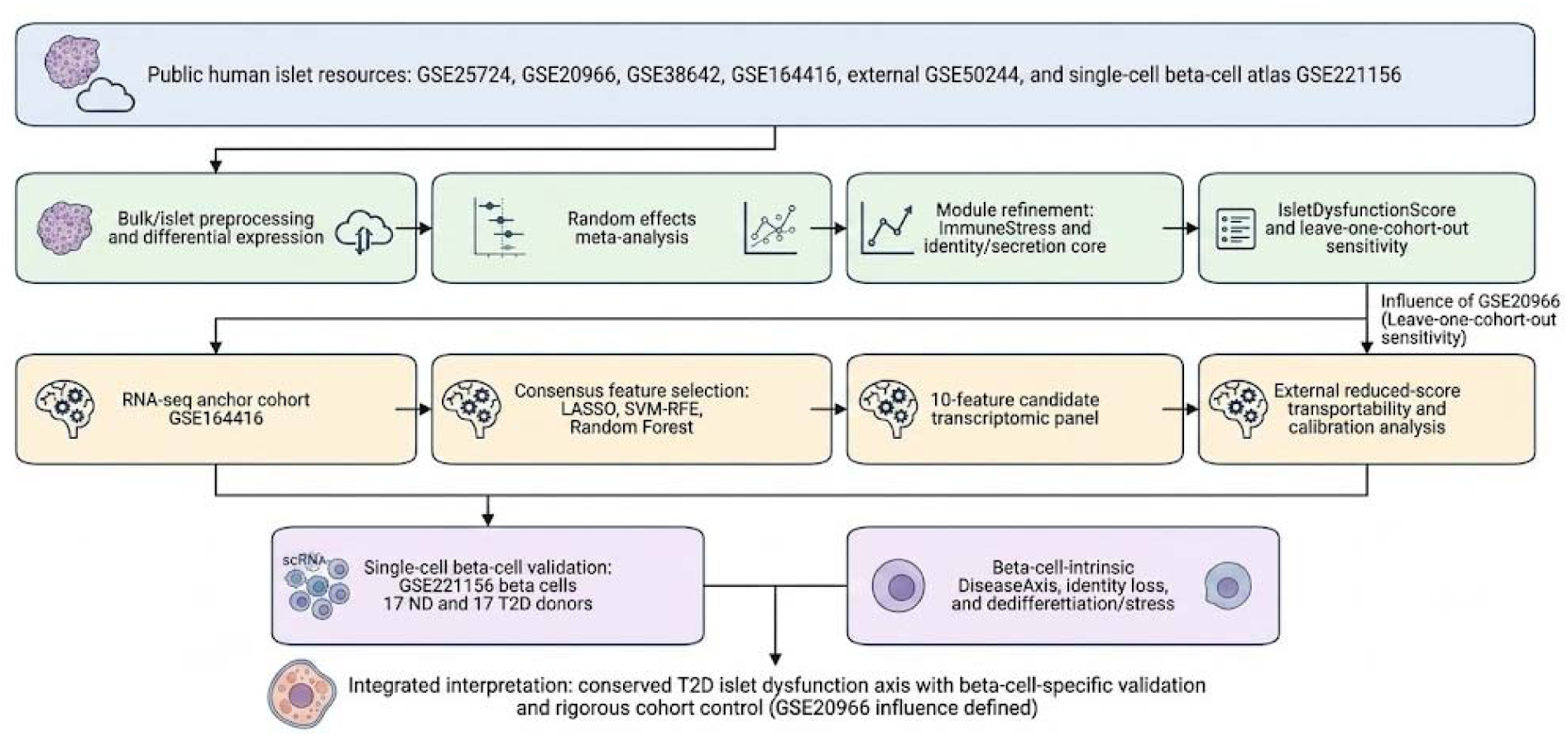
Study overview. The workflow integrates cross-platform bulk/islet meta-analysis, module refinement, candidate-panel machine-learning discovery, external reduced-score validation, and independent single-cell beta-cell validation. *Alt text: Flowchart of the study design showing four bulk or beta-cell-enriched discovery cohorts, meta-analysis and module refinement, machine-learning candidate-panel development, external bulk validation, and independent GSE221156 single-cell beta-cell validation*.

## 2. MATERIALS AND METHODS

### 2.1 Study Design and Public Cohorts

The primary discovery and cross-platform integration arm reanalyzed four public human pancreatic islet transcriptomic cohorts: GSE25724 (15), GSE20966 (16), GSE38642 (17), and GSE164416 (18) (Table 1). These datasets were selected because they contain human islet or beta-cell-enriched transcriptomic profiles with identifiable non-diabetic (ND) and T2D donor groups and gene-level expression profiles suitable for within-cohort differential expression and cross-cohort directionality analysis. The goal was not to privilege any single platform, but to identify signals that persist despite heterogeneous assay and tissue contexts.

**Table 1.** Public cohorts included in the integrative analysis. ND, non-diabetic; T2D, type 2 diabetes; IGT, impaired glucose tolerance; T3cD, type 3c diabetes.

| Cohort | Tissue context | Platform | Samples analysed | Role in study |
| --- | --- | --- | --- | --- |
| GSE25724 | Bulk isolated human islets | Affymetrix microarray | 7 ND / 6 T2D | Cross-platform discovery-support cohort |
| GSE20966 | Laser-capture beta-cell-enriched pancreatic material | Affymetrix microarray | 10 ND / 10 T2D | Endocrine-enriched sensitivity context; not treated as pure beta cells |
| GSE38642 | Bulk human islets | Affymetrix microarray | 54 ND / 9 T2D | Largest legacy bulk-islet replication cohort |
| GSE164416 | Human islets spanning glycaemic continuum | Bulk RNA-seq | 18 ND / 39 T2D (+ IGT, T3cD, other groups) | RNA-seq anchor for stage analysis and ML discovery |
| GSE50244 | Independent bulk human islets | Bulk RNA-seq | 51 ND / 11 T2D labelled subset | External reduced-panel transportability testing |
| GSE221156 beta cells | Reintegrated single-cell beta-cell atlas | 10x 3' v2/v3 scRNA-seq | 17 ND / 17 T2D; 63,350 beta cells after filtering | Independent beta-cell-specific validation |

GSE20966 was retained because it provides beta-cell-enriched laser-capture microdissected material, offering an orthogonal endocrine-enriched context. Here, GSE20966 is explicitly treated as beta-cell-enriched rather than as a pure beta-cell transcriptome. To address this limitation, independent single-cell beta-cell validation was performed using GSE221156.

For the anchor RNA-seq cohort GSE164416, ND and T2D donors were used for binary classification, whereas impaired glucose tolerance (IGT), impaired fasting glucose (IFG), and type 3c diabetes (T3cD) donors were retained only for descriptive glycaemic-stage analyses. The broader HPAP-T2D and IIDP resources are cited as relevant human-islet infrastructure and context for modern donor phenotyping (7–8).

### 2.2 Preprocessing and Differential Expression Analysis

Expression preprocessing was carried out independently for each bulk or microarray dataset. For microarray cohorts, processed expression matrices were imported, probe identifiers were mapped to gene symbols using manufacturer annotation files, duplicated probes were collapsed to a single gene-level profile by retaining the probe with the highest mean signal, and malformed or unmapped identifiers were excluded. Differential expression for microarray datasets was performed using limma with empirical Bayes variance shrinkage (19).

For GSE164416, raw HTSeq gene-level count matrices were obtained from GEO. Counts were log2(x + 1) transformed prior to downstream analyses. Quantile normalization was deliberately excluded from the primary workflow because it can equalize global expression distributions across samples and remove between-group differences that constitute the biological signal (20). Differential expression in the RNA-seq cohort was modelled using DESeq2 for the meta-analysis arm (21) and a limma/voom-like empirical Bayes framework for the ML arm (22). Benjamini-Hochberg false discovery rate correction was used throughout (23).

### 2.3 Gene-Level Integration and Pathway Analysis

Cross-cohort integration was performed using random-effects meta-analysis of study-specific log-fold changes and standard errors implemented in the metafor framework (24). Genes were considered high-confidence meta-signals if they reached nominal significance with directionality preserved in at least three contributing cohorts. Because individual studies differed substantially in power and tissue context, cohort-specific effects were inspected rather than relying exclusively on pooled estimates.

Pathway activity was quantified using sample-wise Gene Set Variation Analysis (GSVA) (25) applied to MSigDB Hallmark gene sets (26). Hallmark results are displayed as a figure rather than only described in text. Pathway-level results were used to contextualize the refined modules, not as primary discovery outputs.

### 2.4 Conserved Module Refinement and Scoring

The two 14-gene modules shown in Table 2 are compact biologically interpretable summaries finalized after inspection of all four discovery cohorts. Consequently, effect sizes computed with these fixed full-data modules in the same four cohorts are treated as descriptive rather than independent validation. The ImmuneStress module comprises MICB, HLA-DRA, HLA-DPA1, IL1R2, IL1RL1, IDO1, SERPING1, FPR3, LTB4R, GBP2, TNFRSF10A, CFH, ADORA3, and APOL1. The ConservedBetaCellIdentitySecretion module comprises RASGRP1, PPP1R1A, ENTPD3, ADCYAP1, FFAR4, TMED6, PLCXD3, PDE8B, CASR, PFKFB2, ACLY, TGFBR3, ASB9, and PPM1K. The latter is a conserved core rather than a comprehensive canonical beta-cell identity set.

**Table 2.**
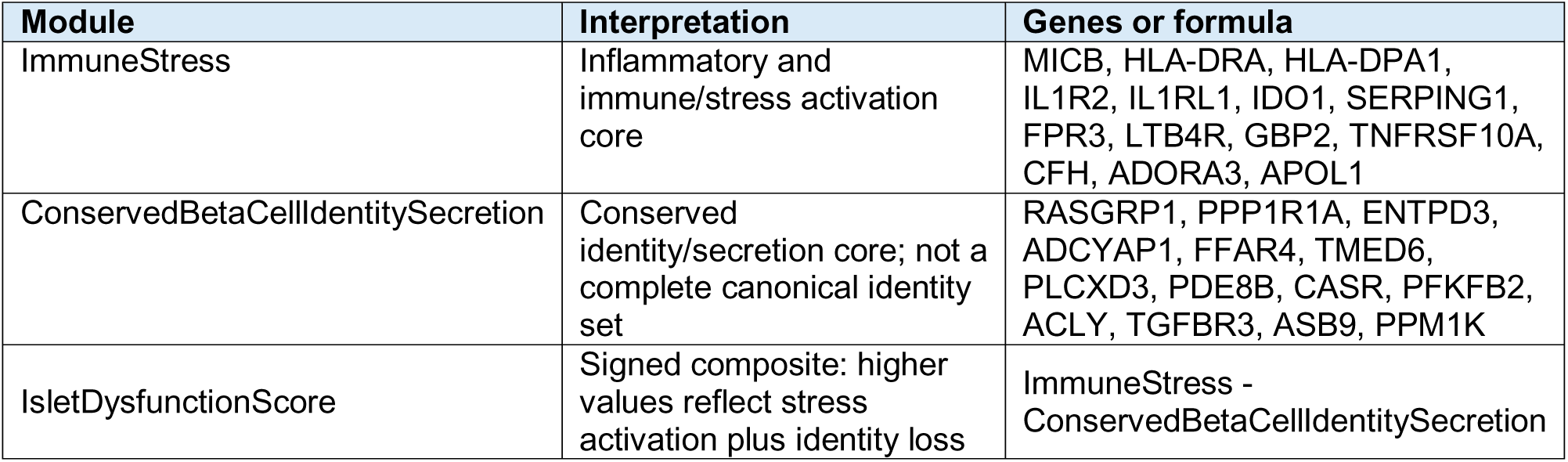
Refined core modules and their biological interpretations. Genes listed in order of meta-analysis effect size magnitude. All genes show directional preservation in beta-cell-enriched tissue (GSE20966).

Within each cohort, expression values were standardized gene-wise across samples. Each fixed module score was computed as the mean standardized expression of member genes. The composite IsletDysfunctionScore was defined as ImmuneStress minus ConservedBetaCellIdentitySecretion, so higher values indicate coordinated immune/stress activation together with loss of beta-cell identity/secretory competence. To obtain out-of-sample module estimates, a separate study-level cross-fitting analysis was performed. For each held-out cohort, de novo 14-gene positive and 14-gene negative modules were selected using only the other three cohorts. Candidate genes had to be technically measurable in the held-out platform, directionally concordant in all three training cohorts, and nominally significant (p < 0.05) in a three-study DerSimonian-Laird random-effects meta-analysis; genes were ranked by absolute meta-analysis z statistic. The held-out disease labels were not used for module selection. Cross-fitted scores were then calculated by the same within-cohort gene standardization and stress-minus-identity subtraction. As a gene-set null, 2000 random sets matched gene-by-gene for mean expression and nonzero detection proportion in the held-out cohort were scored identically; empirical two-sided p values compared the observed absolute effect with this matched null.

### 2.5 Machine Learning Feature Selection and Classification

For the final full-cohort candidate ranking, supervised feature selection was applied to the 184-gene DEG set identified in GSE164416. Three methods were used in parallel: LASSO penalized logistic regression, SVM recursive feature elimination, and Random Forest importance ranking (27). A consensus candidate pool comprised genes selected by at least two methods, and the final full-cohort top-10 ranking was determined by composite importance after internal performance had been estimated by nested resampling. Eight of the final ten features were selected by all three methods in the full-cohort fit and two by two methods.

Primary internal performance estimation used nested stratified 5-fold cross-validation. In each outer training fold, differential-expression filtering was repeated from the full gene matrix using the same thresholds as the discovery workflow (FDR < 0.01 and |log2 fold change| >= 1.5), followed by de novo LASSO, SVM-RFE, Random Forest selection, consensus construction, scaling, and model fitting using training samples only. A soft-voting ensemble of radial-basis-function SVM, Random Forest, L2-penalized Logistic Regression, and Gradient Boosting then predicted the untouched outer fold. Pooled out-of-fold probabilities provided the primary AUC, with a 5000-replicate stratified bootstrap confidence interval; fold-specific AUCs and feature-selection frequencies quantified performance and stability. This design places both DEG filtering and feature selection inside the resampling loop, addressing selection bias in high-dimensional model evaluation (28). Fixed-panel LOOCV and global quantile-normalization analyses are treated only as secondary sensitivity analyses and not as unbiased estimates of classifier performance.

### 2.6 External transportability testing and recalibration sensitivity

External transportability was evaluated in the independent bulk human-islet RNA-seq cohort GSE50244 (29). Annotation differences prevented one-to-one recovery of all final top-10 features, so external testing used a prespecified annotation-compatible 8-gene score drawn from the broader biologically annotated consensus pool: DKK3, PRIMA1, TAFA4, HHATL, PARVG, GABRA2, SLC2A2, and ARG2. PARVG is a consensus-pool gene but is not part of the final full-cohort top-10 ranking; accordingly, this external score is not a literal subset of the final panel and is interpreted as a test of the transportable biological gradient. The signed score was defined as the mean standardized expression of genes upregulated in T2D minus the mean standardized expression of genes downregulated in T2D. Gene-wise standardization parameters and the operating threshold were fixed in discovery and applied unchanged to the external cohort.

To distinguish loss of calibration from loss of discrimination, distribution-shift and recalibration sensitivity analyses compared discovery and external score distributions. Label-free mean-centering and location-scale alignment were interpreted as unsupervised recalibration sensitivity procedures. Externally optimized thresholds or label-based logistic recalibration were interpreted only as descriptive upper bounds (11–14).

### 2.7 Single-cell beta-cell validation using GSE221156

To address beta-cell specificity, we analyzed the reintegrated beta-cell dataset from GSE221156/CZ CELLxGENE, which contains 99,029 beta cells from a 48-donor human islet atlas (9–10). Cells were filtered to ND and T2D donors, yielding 63,350 beta cells from 17 ND and 17 T2D donors. The analysis used donor-balanced subsampling and donor-level aggregation to avoid treating individual cells as independent biological replicates.

Gene identifiers were harmonized to gene symbols when available, while preserving Ensembl identifiers. For single-cell analysis, ImmuneStress used the same 14-gene set as the bulk framework. BetaIdentitySecretion was an expanded beta-cell identity/secretion module comprising the 14-gene conserved core plus SLC2A2, GABRA2, INS, IAPP, PDX1, MAFA, and NKX6-1. To provide a terminology-consistent primary validation, cell-level ImmuneStress and BetaIdentitySecretion scores were standardized across the analyzed ND/T2D beta-cell set and combined as a single-cell analog of IsletDysfunctionScore: z(ImmuneStress) -z(BetaIdentitySecretion). Donor-level means were then compared between ND and T2D using Mann-Whitney and Welch tests. Because the single-cell identity module is expanded and the preprocessing differs from the bulk cohorts, this quantity tests directional and structural transfer of the framework but is not numerically identical to the bulk IsletDysfunctionScore.

Exploratory single-cell modules were analyzed separately and were not treated as interchangeable with IsletDysfunctionScore. DedifferentiationStress comprised ALDH1A3, SOX9, HES1, MYC, LDHA, REST, NEUROG3, CDKN1A, and CDKN2A. ERStress_UPR comprised HSPA5, XBP1, DDIT3, ATF4, ATF6, EIF2AK3, ERN1, HERPUD1, DNAJB9, PDIA4, EDEM1, and SEL1L. OxidativePhosphorylation comprised NDUFA1, NDUFB5, NDUFS1, SDHA, SDHB, UQCRC1, COX4I1, COX5A, ATP5F1A, ATP5F1B, and ATP5MC1.

DiseaseAxis was defined exactly as z(ImmuneStress) + z(ERStress_UPR) + z(DedifferentiationStress) -z(BetaIdentitySecretion). It is an exploratory single-cell composite, is distinct from IsletDysfunctionScore, and is reported only in Supplementary Table S5.

Age, BMI, HbA1c, sex, assay version, and medication annotations were summarized at donor level where available (Supplementary Table S4). Medication metadata in GSE221156 were incomplete: 7 of 17 T2D donors had an antidiabetic medication annotation (five metformin-specific annotations and two nonspecific oral-medication annotations), whereas 10 had no medication annotation. Because no clearly defined untreated T2D reference group was available, drug-specific effects were not estimated.

### 2.8 Biological Annotation and Statistical Analysis

Ensembl gene IDs were mapped to HGNC symbols using Ensembl resources. Gene set enrichment was assessed against KEGG, Gene Ontology Biological Process, and Reactome databases when applicable. Module performance was summarized with cohort-specific group means, standardized effect sizes, two-sample tests, and Wilcoxon/Mann-Whitney tests. In single-cell validation, the donor, not the cell, was the unit of statistical inference. Benjamini-Hochberg correction was applied jointly across the seven donor-level single-cell score tests reported in Table 4 and Supplementary Table S5 (the IsletDysfunctionScore analog plus six component/exploratory scores).

Both female and male donors were represented where sex metadata were available; sex distributions are summarized in Supplementary Table S4. Sex was reported descriptively and was not a primary stratification factor because metadata completeness differed across legacy cohorts.

### 2.9 Ethics Statement

This study analyzed exclusively de-identified publicly available datasets. No new patient recruitment, biological samples, or identifiable data were used. No institutional ethics review or patient consent was required.

## 3. RESULTS

### 3.1 Cohort Characteristics and Dataset Rationale

This study integrates four historical bulk or beta-cell-enriched transcriptomic datasets, one external bulk RNA-seq validation dataset, and one independent beta-cell single-cell validation dataset (Table 1). The four-cohort design tests reproducibility across distinct platforms and tissue contexts. Laser-capture beta-cell-enriched material is not treated as equivalent to purified beta cells. Instead, GSE20966 is interpreted as an endocrine-enriched sensitivity context and GSE221156 provides the direct beta-cell-specific validation layer.

Cohort-specific DEG counts and effect directions were heterogeneous, as expected given differences in platform, sample size, donor inclusion, and tissue context. DEG counts for the primary ND-vs-T2D contrast were GSE25724: 2,135 up and 1,991 down; GSE20966: 34 up and 32 down; GSE38642: 1 up and 1 down; and GSE164416: 1,374 up and 976 down (Figure 2A). The very small DEG yield in GSE38642 and GSE20966 is treated as a limitation of single-cohort differential expression and as a rationale for using direction-aware integration and module-level analysis rather than individual-cohort DEG counts alone.

**Figure 2.**
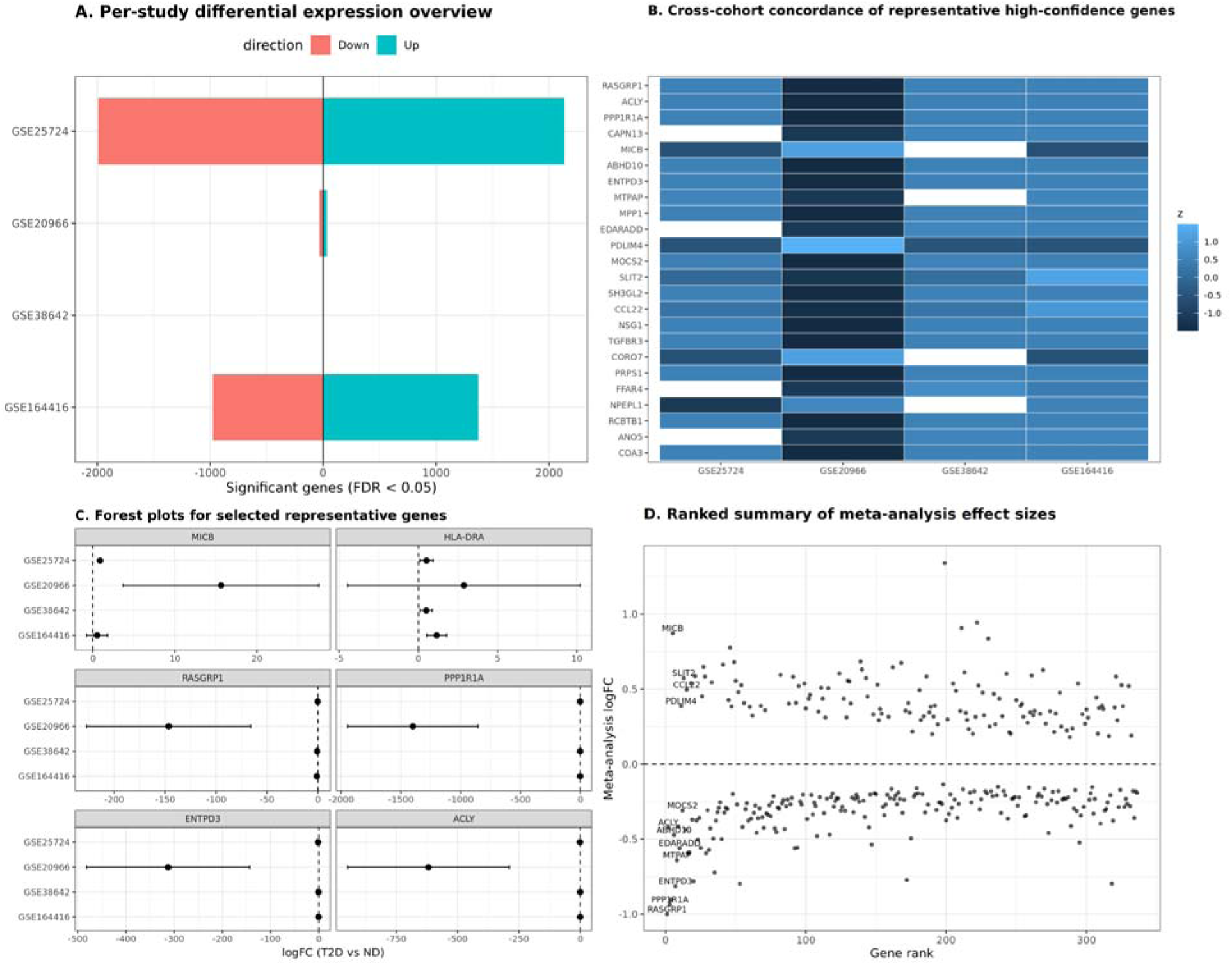
Differential expression and meta-analysis overview. (A) Dataset-specific DEG counts for the ND-vs-T2D contrasts. (B) Representative high-confidence genes across cohorts. (C) Forest plots for selected genes. (D) Ranked meta-analysis effect sizes. *Alt text: Four-panel overview of differential expression and meta-analysis: signed differentially expressed gene counts by cohort, a heatmap of representative high-confidence genes, forest plots for selected genes, and ranked pooled meta-analysis effect sizes*.

### 3.2 Gene-Level Meta-Analysis Identifies a Conserved Transcriptional Core

Random-effects gene-level meta-analysis identified a high-confidence T2D-associated transcriptional core with strong directional preservation across contributing studies (Figure 2B-D). In the beta-cell-enriched GSE20966 context, 334 of 336 meta-analysis genes were detected and 321 of 334 detected genes (96.1%) preserved directionality. This result is interpreted cautiously: it supports compatibility with an endocrine-enriched compartment but does not establish pure beta-cell specificity. The independent GSE221156 beta-cell validation below provides the direct cell-type-specific test.

MICB remains a recurrent positive signal, but its biological interpretation is cautious: it is not used alone as evidence for immune activation. Instead, immune/stress activation is inferred from the combined behavior of the ImmuneStress module and the Hallmark pathway layer.

### 3.3 Hallmark pathway analysis supports inflammatory/stress activation and beta-cell identity erosion

Hallmark GSVA analysis corroborated the gene-level results at the pathway level. The most reproducibly altered pathways included HALLMARK_ALLOGRAFT_REJECTION (mean logFC = 0.037, direction consistency = 1.00, significant in three cohorts), HALLMARK_IL6_JAK_STAT3_SIGNALING (mean logFC = 0.046, direction consistency = 1.00, significant in two cohorts), HALLMARK_INFLAMMATORY_RESPONSE (mean logFC = 0.037, direction consistency = 1.00, significant in two cohorts), HALLMARK_PANCREAS_BETA_CELLS (mean logFC = −0.064, direction consistency = 1.00, significant in two cohorts), and HALLMARK_OXIDATIVE_PHOSPHORYLATION (mean logFC = −0.038, direction consistency = 1.00, significant in two cohorts). These results are shown in Figure 3. Pathway effects were more heterogeneous than the refined module scores, consistent with the broader biological content of Hallmark gene sets.

**Figure 3.**
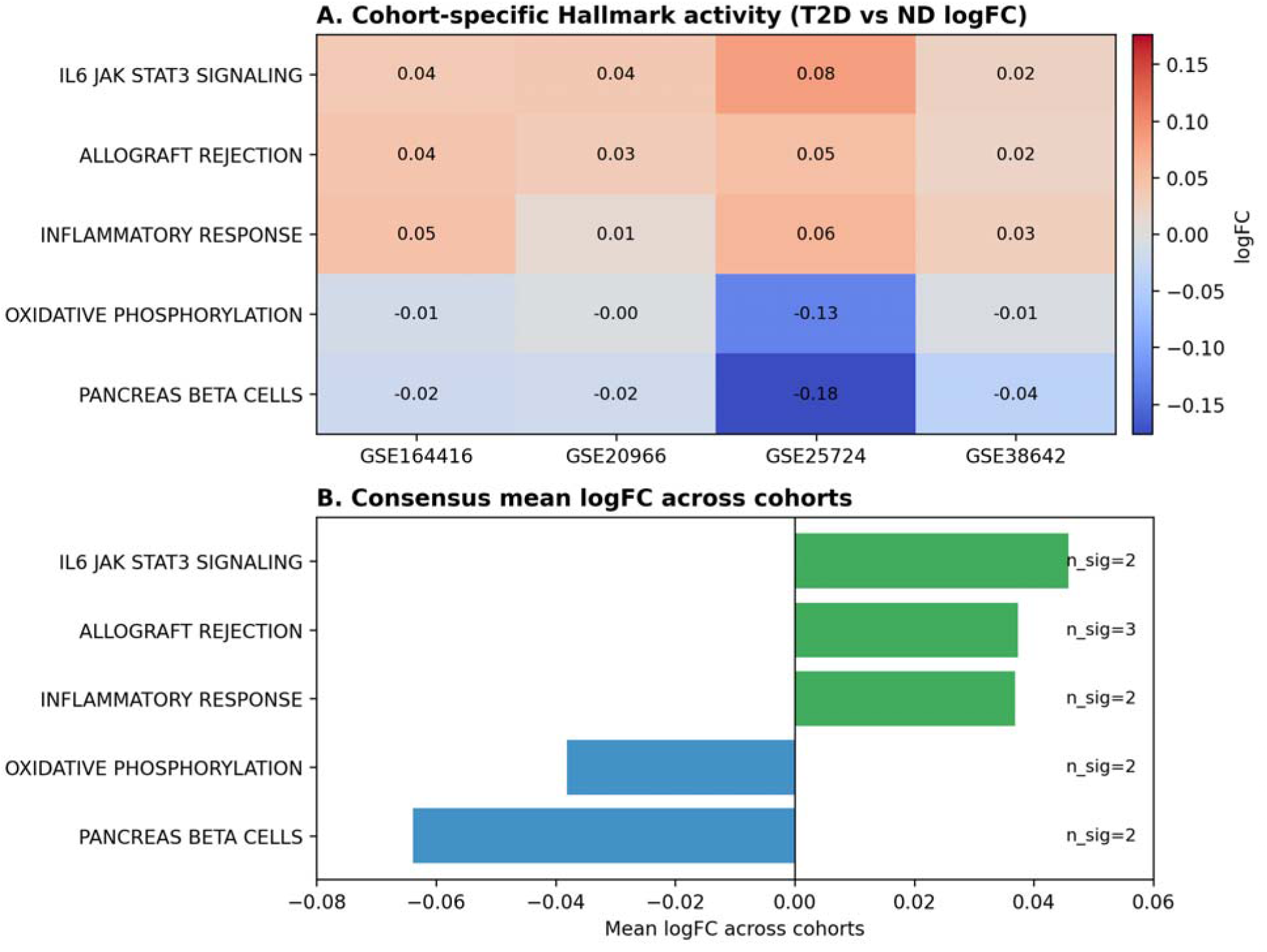
Hallmark pathway activity across cohorts. (A) Cohort-specific Hallmark log-fold changes for reproducible pathways. (B) Consensus mean log-fold change, number of significant cohorts, and direction consistency. Inflammatory/stress-related pathways tended to increase, whereas pancreas beta-cell and oxidative-phosphorylation signatures tended to decrease. *Alt text: Two-panel Hallmark pathway summary showing cohort-specific pathway log-fold changes in a heatmap and consensus mean effects in a horizontal bar plot; inflammatory pathways are positive whereas pancreas beta-cell and oxidative-phosphorylation signatures are negative*.

### 3.4 Refined core modules define a two-axis islet dysfunction framework

The refined module framework distilled the integrated signal into ImmuneStress and ConservedBetaCellIdentitySecretion cores (Table 2; Figure 4). The latter is deliberately interpreted as a conserved cross-cohort core, not as an exhaustive beta-cell identity program. This interpretation prevents the compact module from being confused with a comprehensive canonical beta-cell identity gene set. Broader canonical beta-cell identity genes are treated as complementary validation targets rather than as members of the compact conserved core.

**Figure 4.**
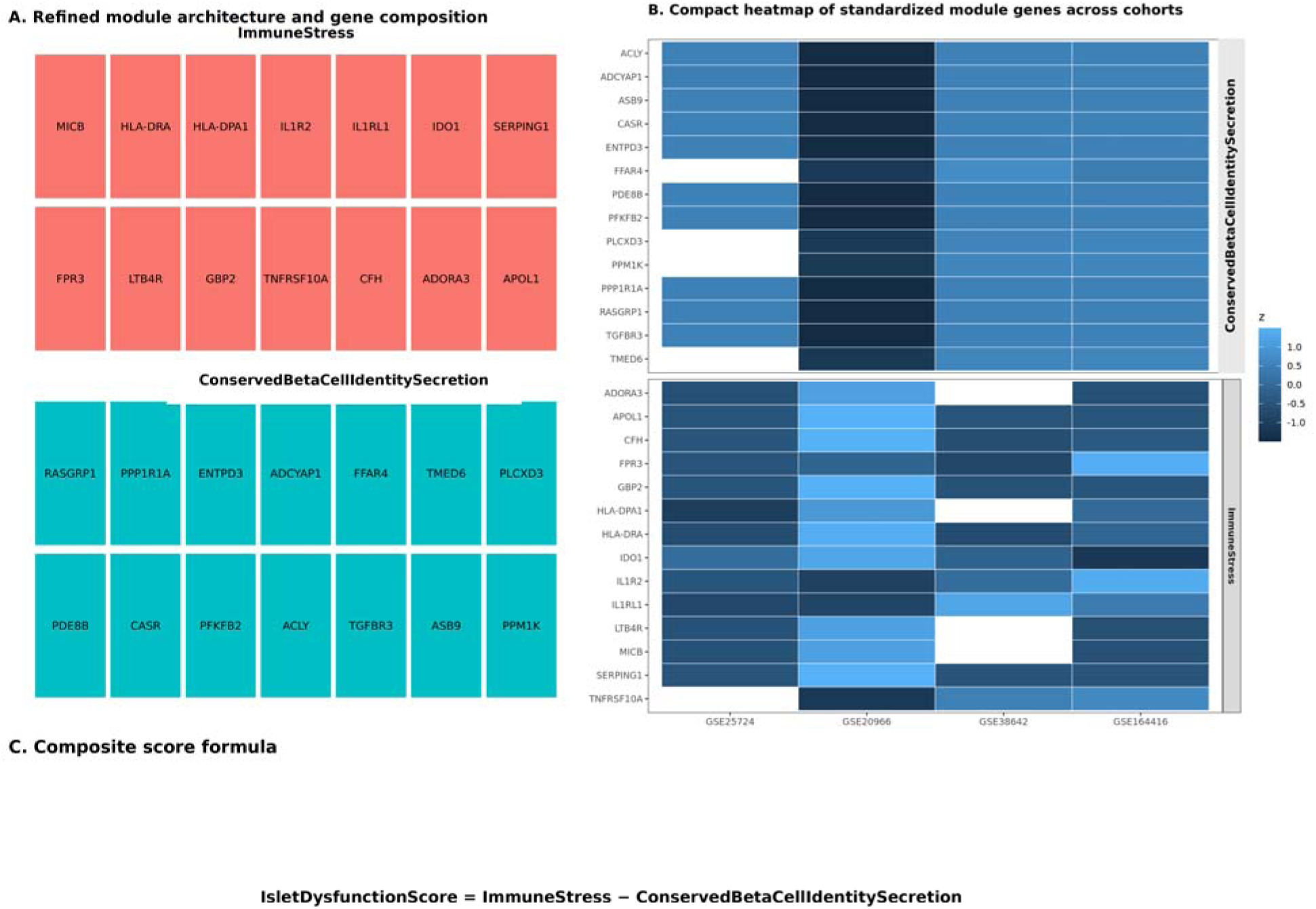
Refined core modules. (A) ImmuneStress and ConservedBetaCellIdentitySecretion modules with gene membership. (B) Within-cohort standardized expression heatmap. (C) IsletDysfunctionScore formula and rationale. *Alt text: Three-panel depiction of the refined ImmuneStress and ConservedBetaCellIdentitySecretion modules, a cross-cohort standardized-expression heatmap, and the IsletDysfunctionScore formula combining stress activation with identity loss*.

The IsletDysfunctionScore was defined as ImmuneStress minus ConservedBetaCellIdentitySecretion. This score is interpretable because higher values represent a coordinated increase in immune/stress activation together with loss of beta-cell identity/secretory competence. The score is used as an integrative summary, while avoiding the implication that the arithmetic form alone establishes mechanism.

### 3.5 Descriptive module effects and study-level cross-fitted validation

Using the fixed full-data modules, descriptive IsletDysfunctionScore effects were positive in GSE25724 (d = 2.18), GSE20966 (d = 2.74), GSE38642 (d = 1.69), and GSE164416 (d = 1.63; Figure 5; Supplementary Table S1 (30)). Descriptive random-effects pooling gave Hedges g = 1.84 (95% CI 1.40-2.27, I2 = 0%; Figure 6; Supplementary Table S2 (30)), whereas the two components were more heterogeneous (ImmuneStress I2 = 70.6%; ConservedBetaCellIdentitySecretion I2 = 77.6%). The lower heterogeneity of the difference score is compatible with partial cancellation of cohort-specific deviations in its two correlated components and should not be interpreted as evidence that the underlying biology is identical across cohorts.

**Figure 5.**
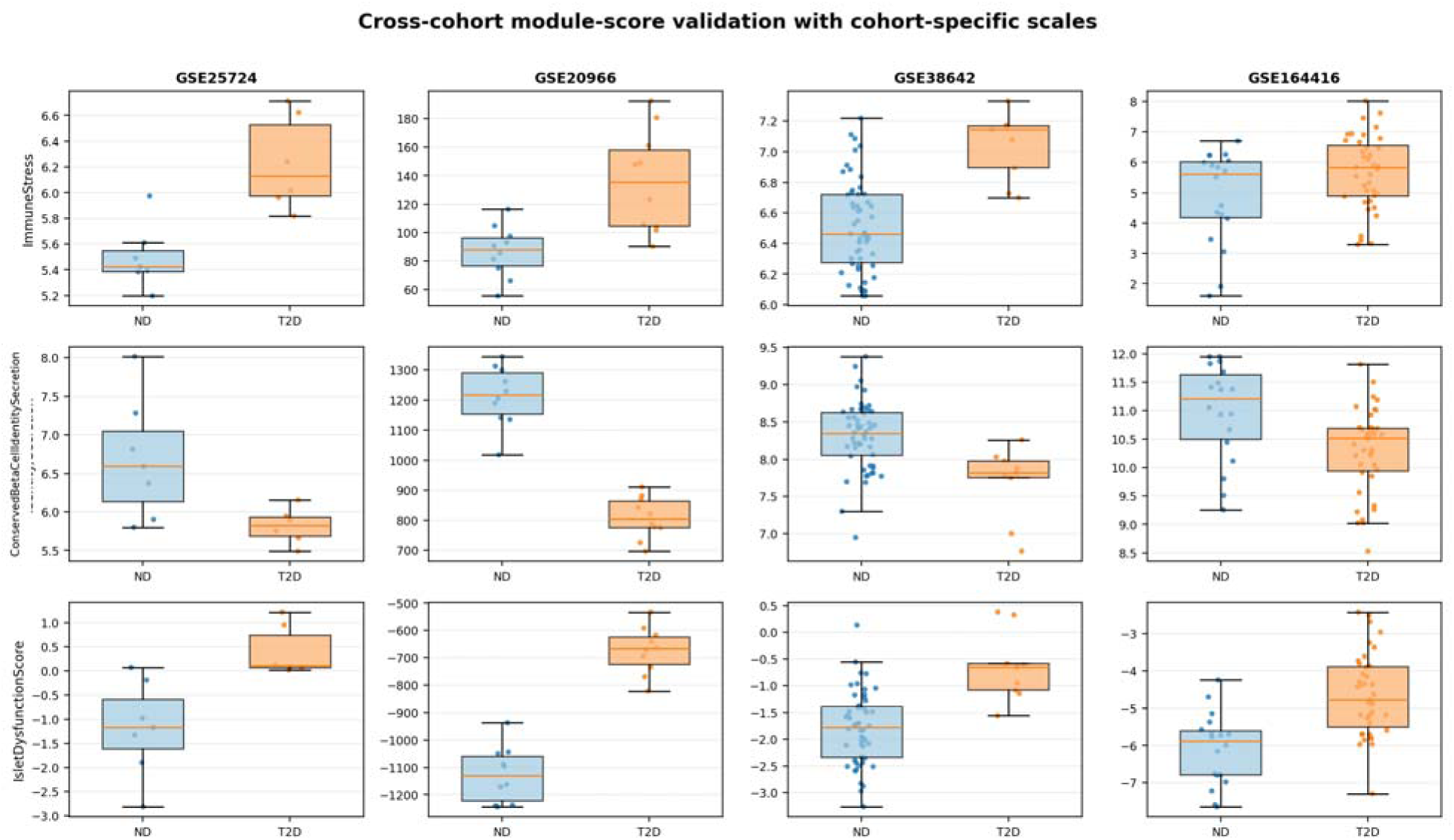
Descriptive full-data module-score distributions. Donor/sample-level scores for the fixed modules are shown separately for each cohort with cohort-specific scales. Because module membership was finalized using information from all four discovery cohorts, these distributions are descriptive; independent generalization is evaluated by study-level cross-fitting in Supplementary Table S6. *Alt text: Grid of donor-or sample-level boxplots comparing non-diabetic and T2D module scores across GSE25724, GSE20966, GSE38642, and GSE164416 using cohort-specific vertical scales*.

**Figure 6.**
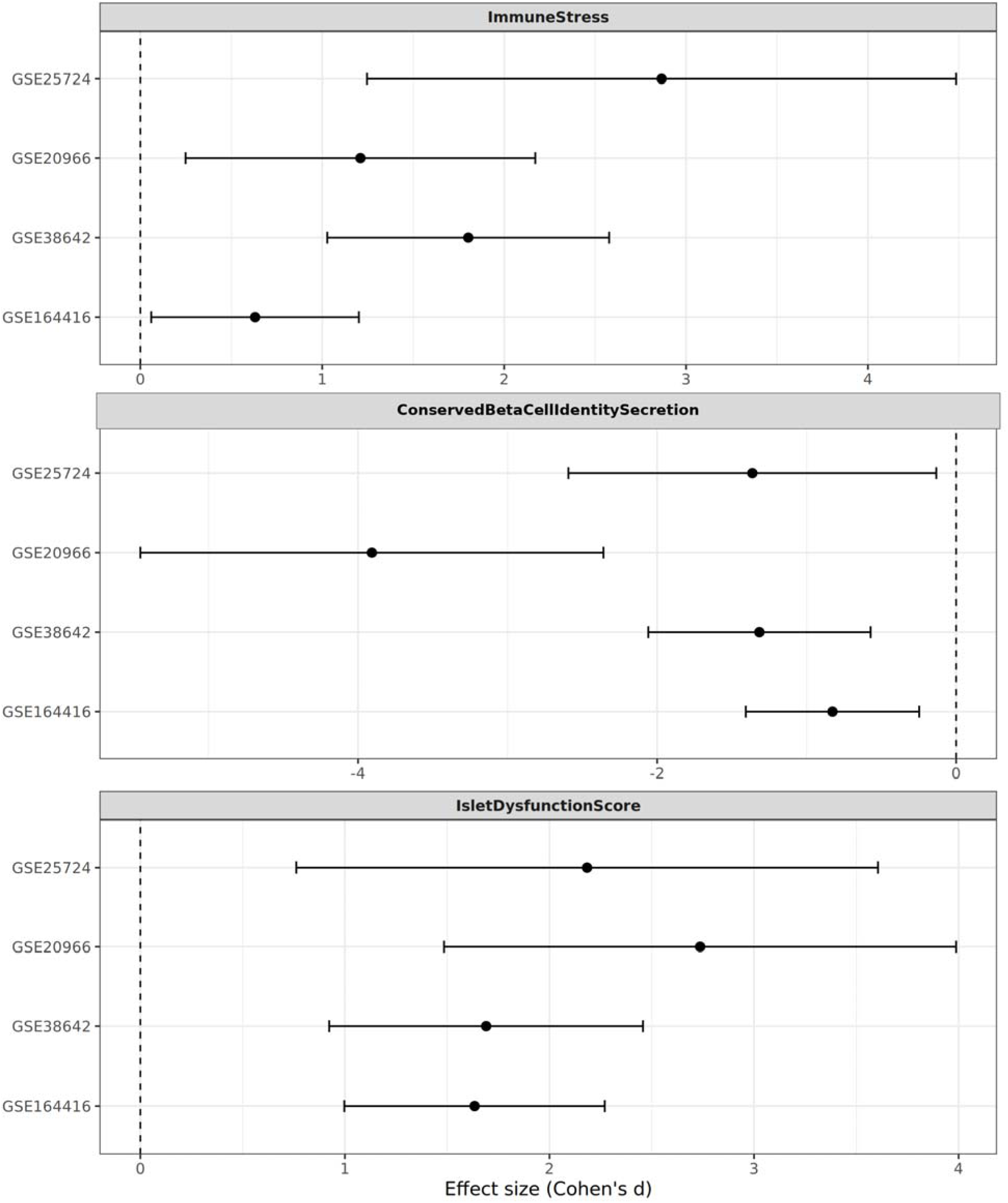
Descriptive full-data forest plots of module effects. Per-cohort standardized effect sizes and random-effects pooled estimates are shown for the fixed ImmuneStress, ConservedBetaCellIdentitySecretion, and IsletDysfunctionScore modules. These full-data estimates are complemented by study-level cross-fitted held-out effects in Supplementary Table S6. *Alt text: Three stacked forest plots showing cohort-specific effect sizes and confidence intervals for ImmuneStress, ConservedBetaCellIdentitySecretion, and IsletDysfunctionScore across the four discovery cohorts*.

Study-level cross-fitting provided the independent generalization test because module membership was re-derived without the held-out cohort. The cross-fitted IsletDysfunctionScore remained positive in all four held-out datasets: GSE25724 d = 1.01, GSE20966 d = 1.36, GSE38642 d = 0.39, and GSE164416 d = 1.13. Random-effects pooling of the four out-of-sample estimates gave Hedges g = 0.905 (95% CI 0.501-1.309, p = 1.14 x 10^-5, I2 = 5.5%; Supplementary Table S6 (30)). In the expression-and detection-matched random gene-set null, the composite empirical p values were 0.397, 0.0245, 0.419, and 0.0020, respectively. Thus, the direction generalized to every held-out cohort, but evidence that the magnitude exceeded matched random gene sets was strongest in GSE20966 and GSE164416. In particular, the positive but modest GSE38642 effect provides a more conservative interpretation of this cohort despite its very small number of individually significant DEGs.

### 3.6 Stage-Dependent Behavior across the glycemic continuum

In the anchor RNA-seq cohort GSE164416, the IsletDysfunctionScore followed a coherent disease ordering across ND, intermediate glycaemic states, T3cD, and T2D: ND mean = −0.661, IGT mean = −0.327, T3cD mean = 0.167, and T2D mean = 0.469. A trend model across disease stage showed a positive stage coefficient for IsletDysfunctionScore (estimate = 0.590, p = 9.03 x 10^-9), with decreasing IdentitySecretion (estimate = −0.391, p = 9.87 x 10^-4) and increasing StressInflammation (estimate = 0.199, p = 0.0329). This pattern supports a graded transcriptional reprogramming of islet state rather than a simple binary contrast, but is interpreted as cross-sectional disease-stage association rather than causal longitudinal progression.

### 3.7 Machine Learning identifies a 10-gene candidate transcriptomic panel

The three-method consensus feature-selection pipeline identified a final 10-feature candidate panel in the executable run: GABRA2, ENSG00000284653, PRIMA1, RNU1-70P, SLC2A2, ARG2, DKK3, TAFA4, HHATL, and ENSG00000227589 (Table 3; Figure 7). Eight features were selected by all three methods and two by two methods. Because ENSG00000227589 remains unannotated and because not all non-coding/unannotated features are portable across external resources, the external GSE50244 analysis is presented as a reduced-score validation of the annotation-compatible biological gradient, not as full validation of every final top-10 feature. This is described as a candidate transcriptomic panel rather than a clinically validated diagnostic test. Single-gene AUCs ranged from 0.828 to 0.941 for the final features, while the panel combined complementary information across beta-cell identity loss, glucose transport, GABA signaling, arginine metabolism, WNT/developmental signaling, neuroimmune signaling, RNA processing, and non-coding or unannotated features. The biological annotation and co-expression context of the candidate-panel genes are summarized in Supplementary Figure S1 (31).

**Figure 7.**
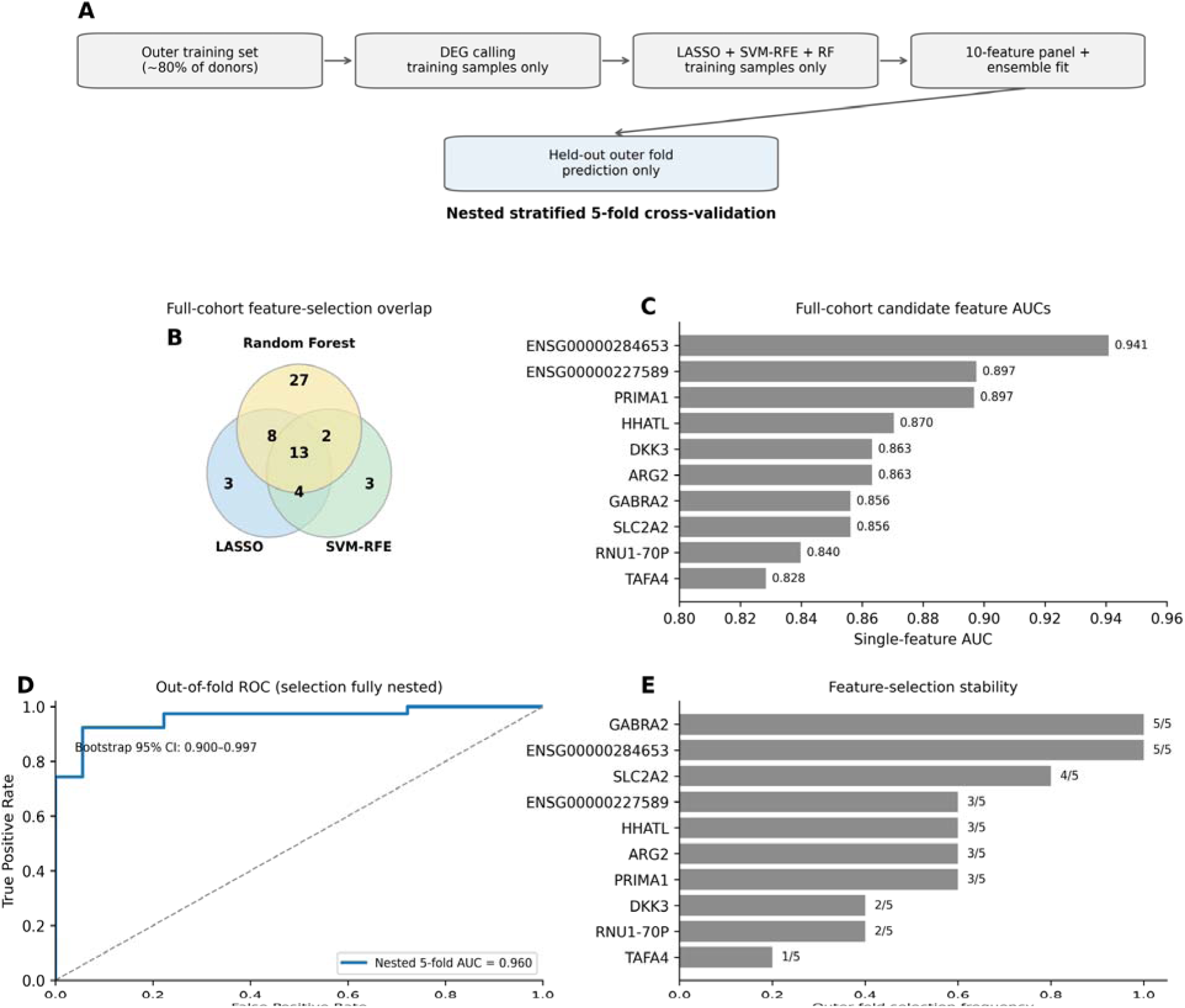
Candidate-panel machine-learning analysis with fully nested validation. (A) Outer-fold workflow in which DEG calling, feature selection, scaling, and model fitting are restricted to each training fold. (B) Full-cohort overlap among LASSO, SVM-RFE, and Random Forest. (C) Full-cohort single-feature AUCs. (D) Pooled outer-fold ROC from nested stratified 5-fold cross-validation (AUC = 0.960; bootstrap 95% CI 0.900-0.997). (E) Selection frequency of the final full-cohort panel genes across outer folds. *Alt text: Five-panel machine-learning summary showing the feature-selection workflow, overlap among LASSO, SVM-RFE and Random Forest, individual feature AUCs, the leave-one-out ROC curve for the ensemble, and a preprocessing/leakage sensitivity ROC comparison*.

**Table 3.** Final 10-feature candidate transcriptomic panel from the full-cohort fit. Eight features were selected by all three methods and two by two methods. The external GSE50244 score is drawn from the broader annotation-compatible consensus pool and includes PARVG, which is not in the final top-10 ranking; it is therefore interpreted separately from validation of the final panel.

| Gene | Biotype | Direction in T2D | Single-gene AUC | Methods selected | Biological context |
| --- | --- | --- | --- | --- | --- |
| GABRA2 | protein-coding | down | 0.856 | 3/3 | GABA-A receptor alpha2 subunit; islet GABA signaling; beta-cell identity context |
| ENSG00000284653 | lncRNA | up | 0.941 | 2/3 | Novel lncRNA; strong upregulated non-coding candidate; functional validation needed |
| PRIMA1 | protein-coding | up | 0.897 | 3/3 | Acetylcholinesterase anchoring; neural-islet signaling context |
| RNU1-70P | pseudogene | down | 0.840 | 3/3 | snRNA pseudogene; RNA processing/splicing candidate |
| SLC2A2 | protein-coding | down | 0.856 | 3/3 | GLUT2 glucose transporter; canonical beta-cell identity marker |
| ARG2 | protein-coding | down | 0.863 | 3/3 | Mitochondrial arginase 2; arginine-NO metabolism and oxidative-stress context |
| DKK3 | protein-coding | up | 0.863 | 3/3 | WNT pathway antagonist; beta-cell dedifferentiation context |
| TAFA4 | protein-coding | up | 0.828 | 3/3 | TAFA chemokine-like family member; neuroimmune signaling context |
| HHATL | protein-coding | up | 0.870 | 2/3 | Hedgehog acyltransferase-like; developmental signaling context |
| ENSG00000227589 | unknown | unknown | 0.897 | 3/3 | Unannotated Ensembl feature; selected by all three methods; manual annotation and validation required |

**Table 4.** Primary donor-level single-cell beta-cell validation in GSE221156. The donor, not the cell, is the unit of inference. BH q values use Benjamini-Hochberg correction across seven donor-level single-cell score tests (the primary analog plus six component/exploratory scores).

| Metric | ND donors | T2D donors | ND mean | T2D mean | Delta T2D-ND | Mann-Whitney p | Welch p | BH q |
| --- | --- | --- | --- | --- | --- | --- | --- | --- |
| IsletDysfunctionScore analog | 17 | 17 | -0.3429 | 0.3703 | 0.7132 | 0.000944 | 0.000309 | 0.00249 |
| BetaIdentitySecretion | 17 | 17 | 0.8792 | 0.7563 | -0.1229 | 0.000572 | 0.000207 | 0.00249 |
| ImmuneStress | 17 | 17 | 0.0052 | 0.0074 | 0.0021 | 0.0388 | 0.0925 | 0.0543 |

Although the compact candidate panel and the refined modules were generated by distinct analytical procedures, their biological convergence on beta-cell identity loss, dedifferentiation-linked signaling, and stress-associated programs is summarized in Supplementary Figure S2 (30,31).

### 3.8 Nested internal validation and feature-selection stability

Fully nested stratified 5-fold cross-validation produced a pooled out-of-fold AUC of 0.960 (bootstrap 95% CI 0.900-0.997), sensitivity 0.923, specificity 0.833, F1 = 0.923, MCC = 0.756, and accuracy = 0.895 (Figure 7D; Supplementary Table S3 (31)). Fold-specific AUCs ranged from 0.917 to 1.000 (mean 0.977, SD 0.036). Feature selection was stable for part, but not all, of the full-cohort panel: GABRA2 and ENSG00000284653 were selected in 5/5 outer folds, SLC2A2 in 4/5, HHATL, ARG2, PRIMA1, and ENSG00000227589 in 3/5, DKK3 and RNU1-70P in 2/5, and TAFA4 in 1/5 (Figure 7E; Supplementary Table S7 (31)). The reduction from the fixed-panel LOOCV estimate of 1.000 to the nested estimate of 0.960 demonstrates the expected optimism when feature selection is performed before resampling; the nested estimate is therefore used as the primary internal performance result.

### 3.9 External transportability reveals preserved discrimination but non-portable thresholds

External testing in GSE50244 retained strong rank-based discrimination with the annotation-compatible 8-gene consensus-pool score (AUC = 0.907; DeLong 95% CI 0.818-0.996) in 62 labelled donors (51 ND and 11 T2D; Figure 8). Directional concordance was observed for six of eight genes (75%): DKK3, PRIMA1, PARVG, GABRA2, SLC2A2, and ARG2 replicated the discovery direction, whereas TAFA4 and HHATL did not. Because PARVG is not in the final top-10 ranking, this analysis tests the broader portable consensus pool rather than a strict reduced subset of the final panel. The frozen discovery threshold failed in the external cohort, yielding sensitivity = 1.000, specificity = 0.000, balanced accuracy = 0.500, and MCC = 0.000 because both external ND and T2D donors were shifted above the discovery cutoff. The score is therefore transportable as a rank-based biological gradient but not as a fixed diagnostic operating threshold.

**Figure 8.**
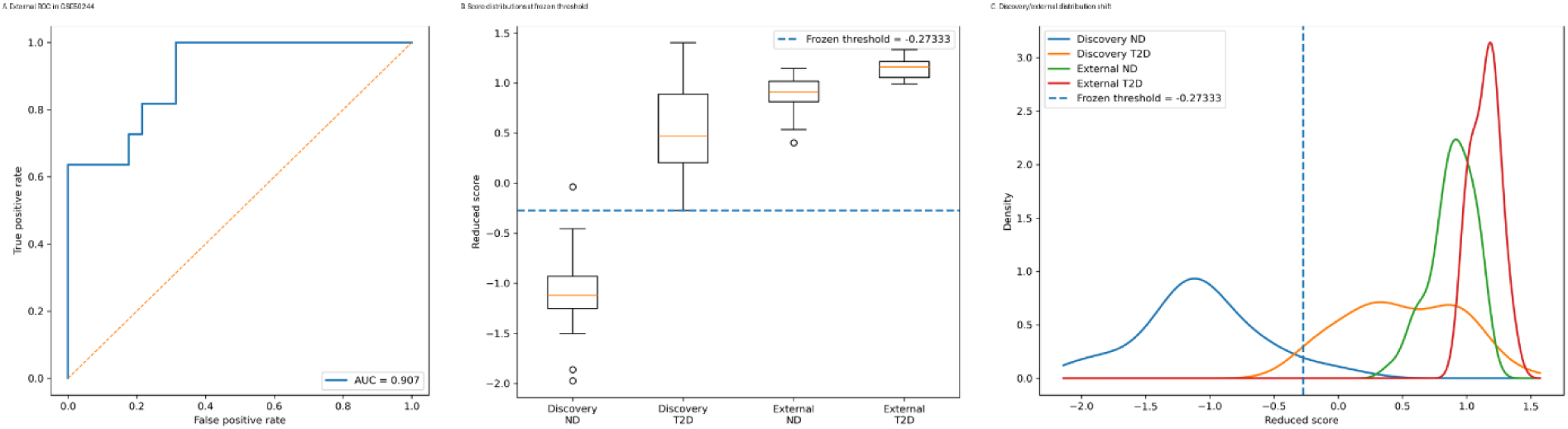
External transportability and calibration-shift analysis. The annotation-compatible 8-gene consensus-pool score retained strong rank-based discrimination in GSE50244 (AUC = 0.907; DeLong 95% CI 0.818-0.996) but failed at the frozen discovery threshold because of cohort-level score shift. The external score is drawn from the broader consensus pool and is not a strict subset of the final top-10 ranking because it includes PARVG. *Alt text: Three-panel external validation summary showing the GSE50244 ROC curve, reduced-score distributions relative to the frozen discovery threshold, and discovery-versus-external score density distributions illustrating calibration shift*.

Distribution-shift and recalibration sensitivity analyses supported calibration drift rather than complete loss of biological signal. Relative to discovery, the external score shifted upward by +0.936 overall, +1.972 in ND donors, and +0.647 in T2D donors. Unsupervised mean-centering modestly improved specificity to 0.137 and balanced accuracy to 0.569, while unsupervised location-scale alignment improved specificity to 0.412 and balanced accuracy to 0.706 without changing the AUC. Label-based external Youden optimization achieved balanced accuracy = 0.843 but is presented only as a descriptive upper bound because it uses external labels. These results are presented as a boundary condition for translational use rather than as evidence of a clinically validated fixed-threshold test.

### 3.10 Covariate sensitivity of the candidate panel

Status-adjusted partial correlations between panel genes and age, BMI, or HbA1c did not survive Benjamini-Hochberg correction across 30 tests (Figure 9A). Residualized fixed-panel LOOCV retained strong discrimination after age/BMI adjustment (AUC = 0.984, specificity = 0.944, MCC = 0.881) and after full age/BMI/HbA1c adjustment (AUC = 0.950; Figure 9B), although specificity and MCC decreased when HbA1c was included. Because the panel was fixed before these residualization analyses, these values are interpreted only as covariate-sensitivity analyses, not as unbiased estimates of predictive performance; the nested 5-fold AUC is the primary internal performance estimate. HbA1c alone was highly discriminative (AUC = 0.966), so HbA1c is treated as a disease-severity variable that partly overlaps with the transcriptomic signal rather than as a simple nuisance covariate.

**Figure 9.**
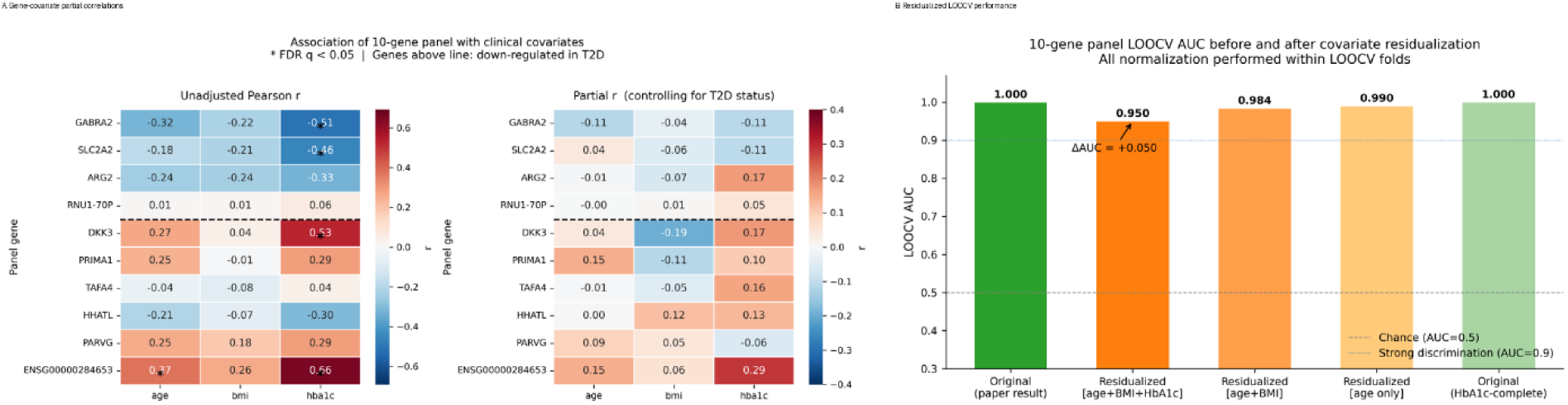
Covariate sensitivity of the candidate panel. (A) Status-adjusted partial correlations showed no FDR-significant gene-covariate associations. (B) Residualized LOOCV showed that age and BMI do not primarily explain the panel, while HbA1c partly overlaps with the disease-severity signal. *Alt text: Covariate-sensitivity figure showing heatmaps of status-adjusted gene associations with age, BMI and HbA1c and a bar chart of leave-one-out AUC after covariate residualization*.

### 3.11 Independent single-cell beta-cell validation of the core dysfunction framework

To test whether the bulk-derived dysfunction framework reflected beta-cell-intrinsic remodeling, we projected the module framework onto the independent GSE221156 beta-cell single-cell atlas. After filtering to ND and T2D donors, the analysis included 63,350 beta cells from 34 donors, with 39,484 ND beta cells and 23,866 T2D beta cells. Donor-level balance was excellent, with 17 ND and 17 T2D donors (Figure 10; Table 4).

**Figure 10.**
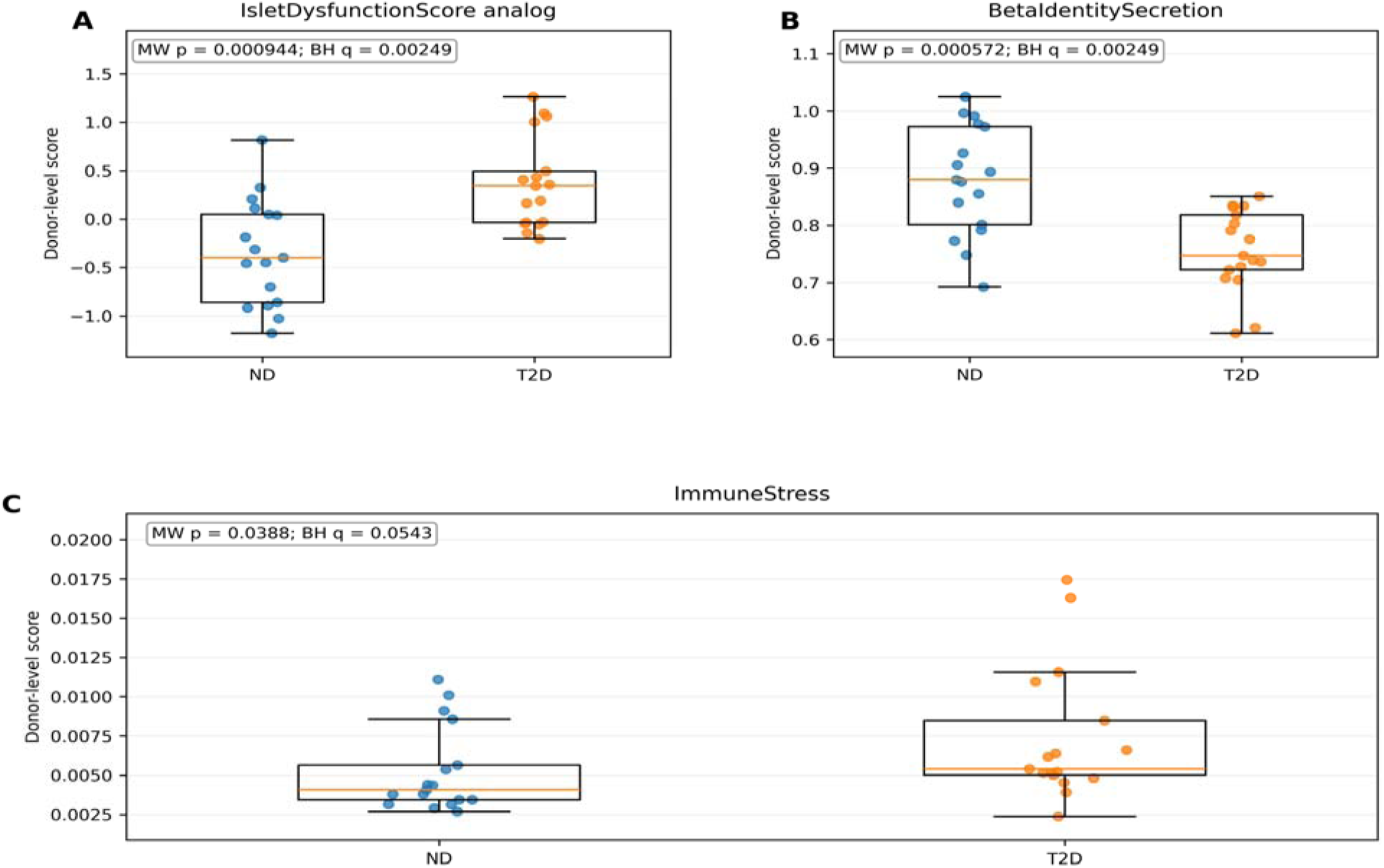
Independent single-cell beta-cell validation using GSE221156. (A) The IsletDysfunctionScore analog is higher in T2D donors (BH q = 0.00249). (B) BetaIdentitySecretion is lower in T2D donors (BH q = 0.00249). (C) ImmuneStress is nominally higher (Mann-Whitney p = 0.0388) but does not survive Benjamini-Hochberg correction across the seven single-cell score tests (q = 0.0543). All comparisons use donor-level means with the donor as the unit of inference. *Alt text: Three donor-level box-and-strip plots comparing non-diabetic and T2D donors in GSE221156 for the single-cell IsletDysfunctionScore analog, BetaIdentitySecretion, and ImmuneStress, with group differences and Mann-Whitney P values displayed outside the data fields*.

The primary single-cell IsletDysfunctionScore analog was higher in T2D donors than in ND donors (ND mean = −0.343, T2D mean = 0.370, Delta = 0.713, Mann-Whitney p = 9.44 x 10^-4, BH q = 0.00249; Welch p = 3.09 x 10^-4; Figure 10A; Table 4). T2D donors also showed lower BetaIdentitySecretion scores (ND mean = 0.879, T2D mean = 0.756, Delta = −0.123, Mann-Whitney p = 5.72 x 10^-4, BH q = 0.00249; Welch p = 2.07 x 10^-4; Figure 10B).

ImmuneStress showed a modest donor-level increase in T2D beta cells (ND mean = 0.00525, T2D mean = 0.00739, Delta = 0.00214; Mann-Whitney p = 0.0388, BH q = 0.0543; Welch p = 0.0925; Figure 10C). Thus, ImmuneStress was nominally different but did not survive correction across the seven single-cell score tests. BetaIdentitySecretion, the IsletDysfunctionScore analog, DedifferentiationStress, and DiseaseAxis remained significant after correction, whereas OxidativePhosphorylation and ERStress_UPR did not (Supplementary Table S5).

Clinical and demographic context is summarized in Supplementary Table S4. In GSE221156, ND and T2D donors had mean ages of 45.4 and 50.4 years, mean BMI values of 28.4 and 33.0 kg/m2, and mean HbA1c values of 5.21% and 7.55%, respectively. Medication information was available for only 7 of 17 T2D donors, all of whom had oral-treatment annotations; the remaining 10 T2D donors lacked a medication annotation. This incomplete and non-random treatment information precludes a reliable estimate of drug-specific transcriptomic effects and is treated as a limitation rather than as evidence of treatment independence.

## 4. DISCUSSION

### 4.1 A Two-Axis Framework with beta-cell-specific validation

The primary contribution of this study is a conservative and validated two-axis framework for human T2D islet transcriptomic dysfunction. The bulk/meta-analysis layer identifies increased immune/stress activation and reduced beta-cell identity/secretory competence. The single-cell validation layer shows that the identity-loss/dedifferentiation component is detectable within beta cells themselves, reducing the concern that the entire signal reflects bulk islet composition.

The laser-capture dataset is informative because it is beta-cell enriched, but it is not pure beta-cell single-cell transcriptomics. Thus, the study does not rely on GSE20966 alone as proof of beta-cell specificity. Instead, GSE20966 is treated as endocrine-enriched support, and GSE221156 provides the direct beta-cell validation.

### 4.2 Interpretation of the single-cell beta-cell validation

The single-cell analysis supports the same directional structure that motivated the bulk IsletDysfunctionScore: loss of beta-cell identity accompanied by increased immune/stress signaling. The primary validation is expressed as a single-cell IsletDysfunctionScore analog together with its two component scores. This analog is not numerically identical to the bulk score because the single-cell BetaIdentitySecretion module includes seven additional canonical beta-cell genes and is calculated after single-cell-specific preprocessing. Accordingly, the result supports transfer of the biological framework rather than universal interchangeability of numerical scores across platforms.

The single-cell results also add nuance. BetaIdentitySecretion showed the strongest component-level shift, whereas ImmuneStress was only nominally increased and did not survive false-discovery-rate correction. Exploratory DedifferentiationStress was higher in T2D, but ERStress_UPR did not increase globally. These findings indicate that stress-related programs are heterogeneous across beta-cell substates and donors and should not be collapsed into a single universal stress phenotype. The exploratory DiseaseAxis is kept separate from the primary IsletDysfunctionScore framework.

### 4.3 Candidate panel as a research transcriptomic signature

The 10-gene panel remains biologically coherent, with genes linked to glucose transport (32), GABA signaling (33), arginine metabolism (34), neural-islet signaling (35), WNT-related dedifferentiation (36), neuroimmune signaling (37), developmental signaling (38), and non-coding regulation (39). Its present value is primarily as a compact research signature for cross-cohort comparison, biological stratification, and prioritization of mechanisms for follow-up. Pancreatic islet or beta-cell transcriptomes are not routinely obtainable in clinical practice, so the current data do not support use of this panel as a direct clinical biomarker. Any future clinical application would require demonstration that related signals can be measured reproducibly in accessible biospecimens, together with prospective validation and platform-aware calibration.

The external GSE50244 analysis is informative as a test of cross-cohort research reproducibility because it separates rank-based discrimination from threshold portability. The reduced annotation-compatible score preserved rank-based separation but failed at the frozen discovery cutoff. This behavior is consistent with calibration drift and reinforces the interpretation of the panel as a research transcriptomic signature rather than a ready-to-deploy diagnostic assay.

### 4.4 Strengths and limitations

This study has several strengths. First, it integrates human islet transcriptomic evidence across multiple public cohorts and assay platforms. Second, study-level cross-fitting separates module definition from held-out evaluation and provides a more conservative assessment of cross-cohort generalizability. Third, machine-learning performance is estimated with fully nested cross-validation in which DEG filtering and feature selection occur inside each outer training fold, while external testing separately evaluates rank-based transportability and calibration. Fourth, donor-level covariate sensitivity analyses assess whether the candidate-panel signal is primarily attributable to measured clinical variables. Finally, independent single-cell beta-cell analysis provides cell-type-resolved support for the identity-loss component of the T2D islet signature.

Several limitations should be considered when interpreting these findings. First, all analyzed cohorts are cross-sectional and samples were obtained after establishment of the recorded glycaemic phenotype. The study therefore cannot determine whether the observed transcriptomic differences precede T2D, contribute causally to disease development, or arise secondarily from chronic hyperglycaemia, insulin resistance, obesity, pharmacological treatment, or other consequences of established disease. The modules should be interpreted as molecular correlates of the T2D islet state rather than as demonstrated initiating mechanisms. Second, donor characteristics differ between disease groups. Age, BMI, and HbA1c were explicitly examined in the principal RNA-seq cohort, and donor-level clinical characteristics for GSE164416 and GSE221156 are summarized in Supplementary Table S4; nevertheless, retrospective public datasets cannot provide complete control of clinical heterogeneity. Third, antidiabetic treatment is a potential modifier of beta-cell transcription. In GSE221156, medication annotations were available for only 7 of 17 T2D donors and all represented oral treatment, leaving no well-defined untreated T2D reference group. Drug-specific effects therefore cannot be separated reliably from disease severity or other donor characteristics. Fourth, GSE20966 provides beta-cell-enriched laser-capture material but not pure beta-cell transcriptomes. Fifth, the refined modules are compact conserved cores rather than exhaustive representations of immune-stress or beta-cell identity biology, and the single-cell identity module is intentionally expanded; numerical scores should therefore not be assumed to be universally interchangeable across platforms. Sixth, the final candidate panel contains non-coding and incompletely annotated features, and external validation was restricted to an annotation-compatible reduced score. Finally, this study evaluates transcriptomic associations only; protein-level validation, functional perturbation, longitudinal studies, and independent prospective cohorts are required before causal or translational claims can be established.

### 4.5 Future Directions

Future work should extend these findings in several directions. Longitudinal or experimentally perturbed human-islet systems are needed to distinguish transcriptomic changes that precede beta-cell failure from those that reflect established metabolic disease. Additional independent single-cell and single-nucleus datasets should test the portability of the core stress/identity framework using harmonized module definitions. Treatment-aware analyses will require cohorts with explicit drug exposure, duration, dose, and untreated comparators. The candidate panel should be evaluated primarily as a research stratification tool and, where suitable tissue resources are available, transcript-level findings should be compared with protein abundance or targeted assays. A clinically relevant extension would require identifying whether components of the islet signature have measurable surrogates in accessible biospecimens rather than assuming that pancreatic tissue transcriptomics can be used routinely.

### 4.6 Conclusions

This study integrates cross-platform human islet transcriptomics, study-level cross-fitted module validation, fully nested machine learning, external bulk validation, and independent single-cell beta-cell analysis. The results support a T2D-associated islet dysfunction framework characterized by increased immune/stress signaling and reduced beta-cell identity/secretory competence. Cross-fitted composite effects were directionally positive in all four held-out bulk cohorts, although matched gene-set null testing showed that effect magnitude was not uniformly exceptional in every cohort. Nested machine learning yielded an internal AUC of 0.960 rather than the optimistic fixed-panel estimate of 1.000, consistent with the external AUC of 0.907. These cross-sectional associations do not establish whether the transcriptional abnormalities are primary drivers or secondary consequences of T2D, and treatment effects cannot be fully resolved from the available metadata. The candidate panel and external consensus-pool score should therefore be viewed as research transcriptomic signatures rather than routine clinical biomarkers.

## Data Availability Statement

All six analyzed datasets are publicly available through the NCBI Gene Expression Omnibus:

GSE25724: https://www.ncbi.nlm.nih.gov/geo/query/acc.cgi?acc=GSE25724; GSE20966: https://www.ncbi.nlm.nih.gov/geo/query/acc.cgi?acc=GSE20966;

GSE38642: https://www.ncbi.nlm.nih.gov/geo/query/acc.cgi?acc=GSE38642; GSE164416: https://www.ncbi.nlm.nih.gov/geo/query/acc.cgi?acc=GSE164416; GSE50244: https://www.ncbi.nlm.nih.gov/geo/query/acc.cgi?acc=GSE50244; GSE221156: https://www.ncbi.nlm.nih.gov/geo/query/acc.cgi?acc=GSE221156.

All analysis scripts are available at the following GitHub repositories: https://github.com/ricardo-romero-ochoa/T2D-islet-integrative

https://github.com/ricardo-romero-ochoa/T2D-islet-ML-panel

https://github.com/ricardo-romero-ochoa/T2D-Islet-Covariate-Sensitivity-Analysis

Single-cell validation code, derived donor-level tables and supplementary material are provided in the GitHub repository:

https://github.com/ricardo-romero-ochoa/scAnalysis-T2D-Islets

and are based on the public GSE221156/CZ CELLxGENE resource.

Reprint requests: Ricardo Romero, Departamento de Ciencias Naturales, Universidad Autonoma Metropolitana, Unidad Cuajimalpa, Avenida Vasco de Quiroga 4871, Col. Santa Fe Cuajimalpa, Alcaldia Cuajimalpa de Morelos, C.P. 05348, Ciudad de Mexico, Mexico.

## Author Contributions

R. Romero: Conceptualization, data curation, formal analysis, methodology, software, visualization, writing, original draft, writing, review and editing.

## REFERENCES

1. International Diabetes Federation. IDF Diabetes Atlas. 10th ed. International Diabetes Federation; 2021. Accessed October 21, 2026. https://diabetesatlas.org

2. American Diabetes Association Professional Practice Committee. Classification and diagnosis of diabetes: Standards of Care in Diabetes-2024. Diabetes Care. 2024;47(suppl 1):S20–S42. doi:10.2337/dc24-S002

3. Spijker HS, Song H, Ellenbroek JH, Roefs MM, Engelse MA, Bos E, Koster AJ, Rabelink TJ, Hansen BC, Clark A, Carlotti F, de Koning EJP. Loss of beta-cell identity occurs in type 2 diabetes and is associated with islet amyloid deposits. Diabetes. 2015;64(8):2928–2938. doi:10.2337/db14-1752

4. Dludla PV, Mabhida SE, Ziqubu K, Nkambule BB, Mazibuko-Mbeje SE, Hanser S, Basson AK, Pheiffer C, Kengne AP. Pancreatic beta-cell dysfunction in type 2 diabetes: implications of inflammation and oxidative stress. World J Diabetes. 2023;14(3):130–146. doi:10.4239/wjd.v14.i3.130

5. Lawlor N, Stitzel ML. (Epi)genomic heterogeneity of pancreatic islet function and failure in type 2 diabetes. Mol Metab. 2019;27(suppl):S15–S24. doi:10.1016/j.molmet.2019.06.002

6. Kaestner KH, Powers AC, Naji A, HPAP Consortium, Atkinson MA. NIH initiative to improve understanding of the pancreas, islet, and autoimmunity in type 1 diabetes: the Human Pancreas Analysis Program. Diabetes. 2019;68(7):1394–1402. doi:10.2337/db19-0058

7. Shapira SN, Naji A, Atkinson MA, Powers AC, Kaestner KH. Understanding islet dysfunction in type 2 diabetes through multidimensional pancreatic phenotyping: the Human Pancreas Analysis Program. Cell Metab. 2022;34(12):1906–1913. doi:10.1016/j.cmet.2022.09.013

8. Brissova M, Niland JC, Cravens J, Olack B, Sowinski J, Evans-Molina C. The Integrated Islet Distribution Program answers the call for improved human islet phenotyping and reporting of human islet characteristics in research articles. Diabetes. 2019;68(7):1363–1365. doi:10.2337/dbi19-0019

9. Bandesh K, Motakis E, Nargund S, Kursawe R, Selvam V, Ansarullah, Bhuiyan RM, Eryilmaz GN, Willett AM, White JK, Krishnan SN, Spracklen CN, Ucar D, Stitzel ML. Deep single-cell decoding of human pancreatic islets reveals T2D beta-cell gene expression defects. EMBO J. 2026;45(11):3978–4005. doi:10.1038/s44318-026-00744-w

10. Motakis E, Nargund S, Kursawe R, Lawlor N, Krishnan SN, Selvam V, Stitzel ML. Dissecting cell-type specific pathophysiological changes in human type 2 diabetic pancreatic islets using single-cell transcriptomic data. Gene Expression Omnibus. GSE221156. Published 2023. Accessed October 21, 2026. https://www.ncbi.nlm.nih.gov/geo/query/acc.cgi?acc=GSE221156

11. Riley RD, Archer L, Snell KIE, Ensor J, Dhiman P, Martin GP, Bonnett LJ, Collins GS. Evaluation of clinical prediction models (part 2): how to undertake an external validation study. BMJ. 2024;384:e074820. doi:10.1136/bmj-2023-074820

12. Van Calster B, McLernon DJ, van Smeden M, Wynants L, Steyerberg EW. Calibration: the Achilles heel of predictive analytics. BMC Med. 2019;17:230. doi:10.1186/s12916-019-1466-7

13. Van Calster B, Steyerberg EW, Wynants L, van Smeden M. There is no such thing as a validated prediction model. BMC Med. 2023;21:70. doi:10.1186/s12916-023-02779-w

14. Steyerberg EW, Borsboom GJ, van Houwelingen HC, Eijkemans MJC, Habbema JDF. Validation and updating of predictive logistic regression models. Stat Med. 2004;23(16):2567–2586. doi:10.1002/sim.1844

15. Dominguez V, Raimondi C, Somanath S, Bugliani M, Loder MK, Edling CE, Divecha N, da Silva-Xavier G, Marselli L, Persaud SJ, Turner MD, Rutter GA, Marchetti P, Falasca M, Maffucci T. Class II phosphoinositide 3-kinase regulates exocytosis of insulin granules in pancreatic beta cells. J Biol Chem. 2011;286(6):4216–4225. doi:10.1074/jbc.M110.200295

16. Marselli L, Thorne J, Dahiya S, Sgroi DC, Sharma A, Bonner-Weir S, Marchetti P, Weir GC. Gene expression profiles of beta-cell enriched tissue obtained by laser capture microdissection from subjects with type 2 diabetes. PLoS One. 2010;5(7):e11499. doi:10.1371/journal.pone.0011499

17. Taneera J, Lang S, Sharma A, Fadista J, Zhou Y, Ahlqvist E, Jonsson A, Lyssenko V, Vikman P, Hansson O, Parikh H, Korsgren O, Soni A, Krus U, Zhang E, Jing XJ, Esguerra JLS, Wollheim CB, Salehi A, Rosengren A, Renstrom E, Groop L. A systems genetics approach identifies genes and pathways for type 2 diabetes in human islets. Cell Metab. 2012;16(1):122–134. doi:10.1016/j.cmet.2012.06.006

18. Wigger L, Barovic M, Brunner AD, Marzetta F, Schoniger E, Mehl F, Kipke N, Friedland D, Burdet F, Kessler C, Lesche M, Thorens B, Bonifacio E, Legido-Quigley C, Barbier Saint Hilaire P, Delerive P, Dahl A, Klose C, Gerl MJ, Simons K, Aust D, Weitz J, Distler M, Schulte AM, Mann M, Ibberson M, Solimena M. Multi-omics profiling of living human pancreatic islet donors reveals heterogeneous beta cell trajectories towards type 2 diabetes. Nat Metab. 2021;3:1017–1031. doi:10.1038/s42255-021-00420-9

19. Ritchie ME, Phipson B, Wu D, Hu Y, Law CW, Shi W, Smyth GK. limma powers differential expression analyses for RNA-sequencing and microarray studies. Nucleic Acids Res. 2015;43(7):e47. doi:10.1093/nar/gkv007

20. Bullard JH, Purdom E, Hansen KD, Dudoit S. Evaluation of statistical methods for normalization and differential expression in mRNA-seq experiments. BMC Bioinformatics. 2010;11:94. doi:10.1186/1471-2105-11-94

21. Love MI, Huber W, Anders S. Moderated estimation of fold change and dispersion for RNA-seq data with DESeq2. Genome Biol. 2014;15:550. doi:10.1186/s13059-014-0550-8

22. Law CW, Chen Y, Shi W, Smyth GK. voom: precision weights unlock linear model analysis tools for RNA-seq read counts. Genome Biol. 2014;15:R29. doi:10.1186/gb-2014-15-2-r29

23. Benjamini Y, Hochberg Y. Controlling the false discovery rate: a practical and powerful approach to multiple testing. J R Stat Soc Series B Stat Methodol. 1995;57(1):289–300. doi:10.1111/j.2517-6161.1995.tb02031.x

24. Viechtbauer W. Conducting meta-analyses in R with the metafor package. J Stat Softw. 2010;36(3):1–48. doi:10.18637/jss.v036.i03

25. Hanzelmann S, Castelo R, Guinney J. GSVA: gene set variation analysis for microarray and RNA-seq data. BMC Bioinformatics. 2013;14:7. doi:10.1186/1471-2105-14-7

26. Liberzon A, Birger C, Thorvaldsdottir H, Ghandi M, Mesirov JP, Tamayo P. The Molecular Signatures Database Hallmark gene set collection. Cell Syst. 2015;1(6):417–425. doi:10.1016/j.cels.2015.12.004

27. Pedregosa F, Varoquaux G, Gramfort A, Michel V, Thirion B, Grisel O, Blondel M, Prettenhofer P, Weiss R, Dubourg V, Vanderplas J, Passos A, Cournapeau D, Brucher M, Perrot M, Duchesnay E. Scikit-learn: machine learning in Python. J Mach Learn Res. 2011;12:2825–2830.

28. Varma S, Simon R. Bias in error estimation when using cross-validation for model selection. BMC Bioinformatics. 2006;7:91. doi:10.1186/1471-2105-7-91

29. Fadista J, Vikman P, Laakso EO, Guerra Mollet I, Esguerra JL, Taneera J, Storm P, Osmark P, Ladenvall C, Prasad RB, Hansson KB, Finotello F, Uvebrant K, Ofori JK, Di Camillo B, Krus U, Cilio CM, Hansson O, Eliasson L, Rosengren AH, Renstrom E, Wollheim CB, Groop L. Global genomic and transcriptomic analysis of human pancreatic islets reveals novel genes influencing glucose metabolism. Proc Natl Acad Sci U S A. 2014;111(38):13924–13929. doi:10.1073/pnas.1402665111

30. Romero R. T2D-islet-integrative. Version 1.0.0. Zenodo. 2026. doi:10.5281/zenodo.20547703

31. Romero R. T2D-islet-ML-panel. Version 1.0.1. Zenodo. 2026. doi:10.5281/zenodo.20549014

32. Thorens B. GLUT2, glucose sensing and glucose homeostasis. Diabetologia. 2015;58(2):221–232. doi:10.1007/s00125-014-3451-1

33. Soltani N, Qiu H, Aleksic M, Glinka Y, Zhao F, Liu R, Li Y, Zhang N, Chakrabarti R, Ng T, Jin T, Zhang H, Lu WY, Feng ZP, Prud’homme GJ, Wang Q. GABA exerts protective and regenerative effects on islet beta cells and reverses diabetes. Proc Natl Acad Sci U S A. 2011;108(28):11692–11697. doi:10.1073/pnas.1102715108

34. Romero MJ, Platt DH, Tawfik HE, Labazi M, El-Remessy AB, Bartoli M, Caldwell RB, Caldwell RW. Diabetes-induced coronary vascular dysfunction involves increased arginase activity. Circ Res. 2008;102(1):95–102. doi:10.1161/CIRCRESAHA.107.155028

35. Perrier NA, Kherif S, Perrier AL, Dumas S, Mallet J, Massoulie J. Expression of PRiMA in the mouse brain. Eur J Neurosci. 2003;18(7):1837–1847. doi:10.1046/j.1460-9568.2003.02914.x

36. Rulifson IC, Karnik SK, Heiser PW, ten Berge D, Chen H, Gu X, Taketo MM, Nusse R, Hebrok M, Kim SK. Wnt signaling regulates pancreatic beta cell proliferation. Proc Natl Acad Sci U S A. 2007;104(15):6247–6252. doi:10.1073/pnas.0701509104

37. Delfini MC, Mantilleri A, Gaillard S, Hao J, Reynders A, Malapert P, Alonso S, Francois A, Barrere C, Seal R, Landry M, Eschallier A, Alloui A, Bourinet E, Delmas P, Le Feuvre Y, Moqrich A. TAFA4, a chemokine-like protein, modulates injury-induced mechanical and chemical pain hypersensitivity in mice. Cell Rep. 2013;5(2):378–388. doi:10.1016/j.celrep.2013.09.013

38. Lucas C, Ferreira C, Cazzanelli G, Franco-Duarte R, Tulha J. Yeast Gup1 proteins are homologues of HHAT(L). J Dev Biol. 2016;4(4):33. doi:10.3390/jdb4040033

39. Akerman I, Tu Z, Beucher A, Rolando DMY, Sauty-Colace C, Benazra M, Nakic N, Yang J, Wang H, Pasquali L, Moran I, Garcia-Hurtado J, Castro N, Gonzalez-Franco R, Stewart AF, Bonner C, Piemonti L, Berney T, Groop L, Kerr-Conte J, Pattou F, Argmann C, Schadt E, Ravassard P, Ferrer J. Human pancreatic beta cell lncRNAs control cell-specific regulatory networks. Cell Metab. 2017;25(2):400–411. doi:10.1016/j.cmet.2016.11.016

