## Supplementary_Material for "Human Islet Transcriptomics in Type 2 Diabetes Reveals Conserved Dysfunction Axes and a Candidate Gene Panel"

**Supplementary figures**

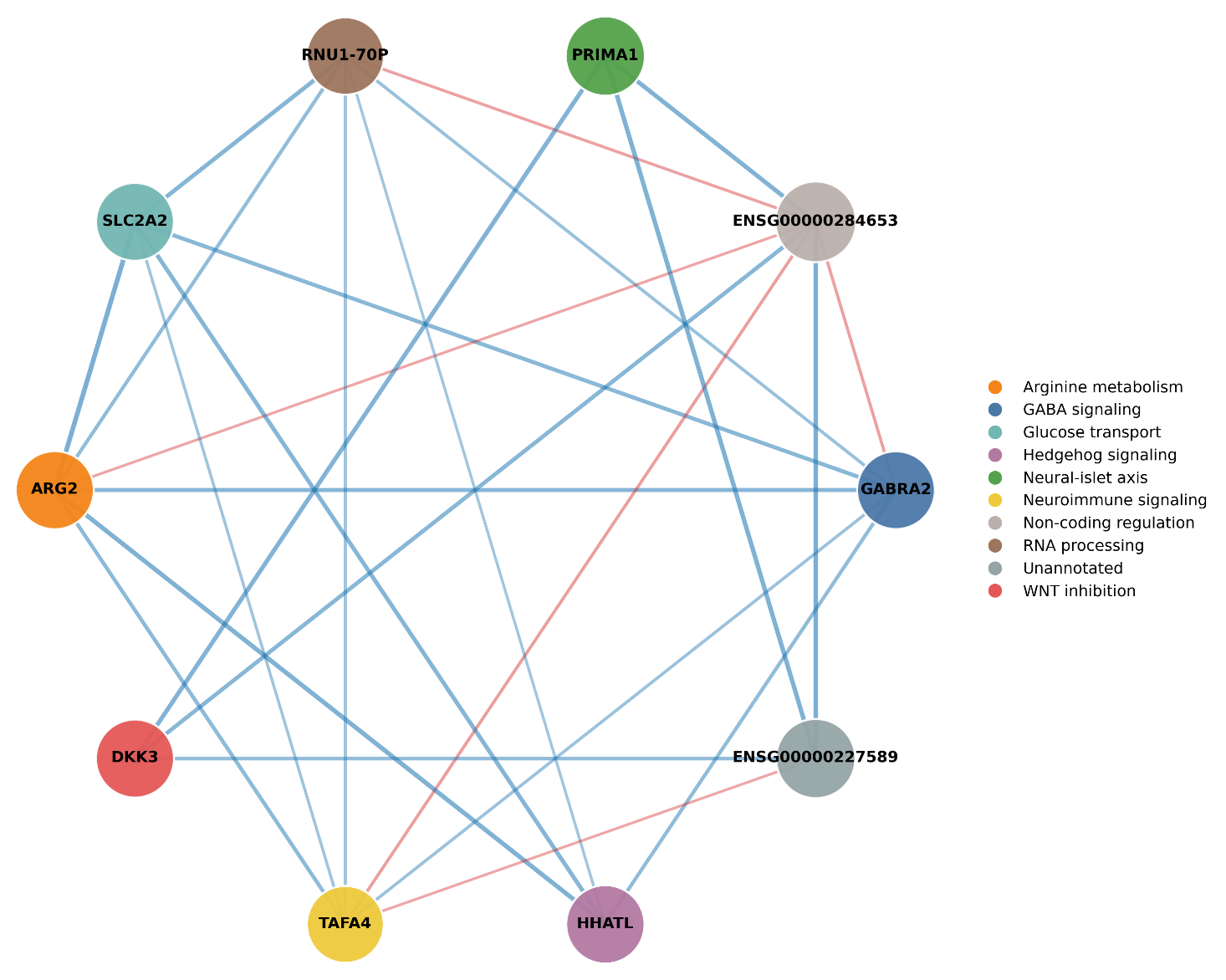

**Supplementary Figure S1.** Biological annotation and pathway context of the 10-feature candidate transcriptomic panel. Network diagram showing candidate-panel features annotated by biological pathway membership, with node size proportional to individual AUC and edge weights reflecting co-expression in the GSE164416 cohort.

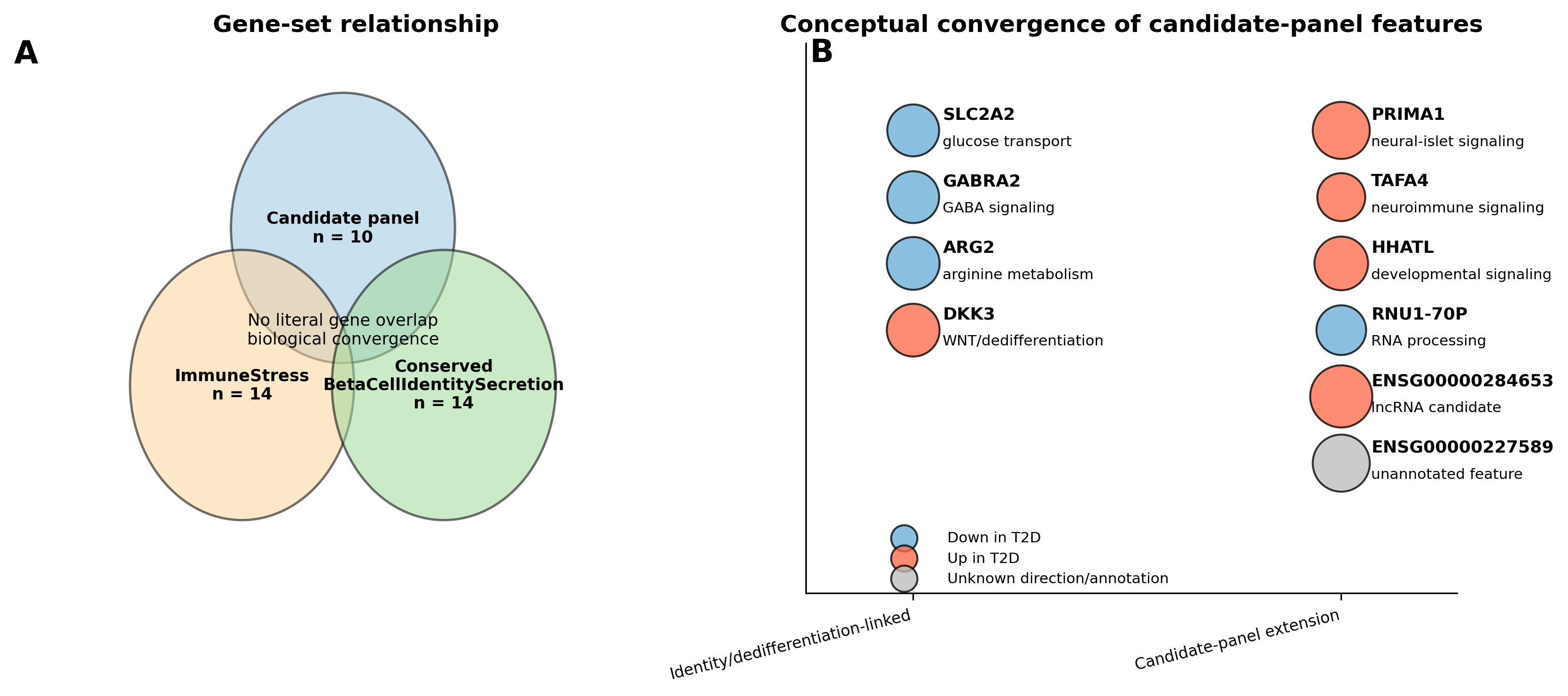

**Supplementary Figure S2.** Convergence between the meta-analysis and machine-learning frameworks. (A) Gene-set relationship among the candidate panel, ImmuneStress module, and ConservedBetaCellIdentitySecretion module. (B) Conceptual organization of candidate-panel features by direction of change and biological context, highlighting convergence on beta-cell identity loss, dedifferentiation-linked signaling, and stress-associated programs.

**Supplementary tables**

Supplementary Table S1. Descriptive full-data cohort-specific effect sizes for the fixed module framework.

| **Cohort** | **n ND** | **n T2D** | **ImmuneStress d** | **ConservedBetaCell IdentitySecretion d** | **IsletDysfunctionScore d** |
| --- | --- | --- | --- | --- | --- |
| GSE25724 | 7 | 6 | 2.87 | -1.36 | 2.18 |
| GSE20966 | 10 | 10 | 1.21 | -3.91 | 2.74 |
| GSE38642 | 54 | 9 | 1.80 | -1.32 | 1.69 |
| GSE164416 | 18 | 39 | 0.63 | -0.83 | 1.63 |

*d, Cohen's d (positive values = higher in T2D; negative values = lower in T2D).*

Supplementary Table S2. Descriptive full-data random-effects pooled standardized effects for the fixed modules.

| **Module / Score** | **Pooled Hedges' g** | **95% CI** | **p value** | **I2 (%)** |
| --- | --- | --- | --- | --- |
| ImmuneStress | 1.45 | 0.62 to 2.27 | 5.9 x 10^-4 | 70.6 |
| ConservedBetaCell IdentitySecretion | -1.66 | -2.64 to -0.67 | 9.7 x 10^-4 | 77.6 |
| IsletDysfunctionScore | 1.84 | 1.40 to 2.27 | <2.3 x 10^-16 | 0.0 |

*Random-effects pooled using a random-effects meta-analysis model. I2, between-cohort heterogeneity percentage; CI, confidence interval.*

Supplementary Table S3. Fully nested stratified 5-fold cross-validation performance of the candidate-panel workflow.

| **Validation scheme** | **AUC (95% CI)** | **F1** | **Sensitivity** | **Specificity** | **MCC** |
| --- | --- | --- | --- | --- | --- |
| Fully nested 5-fold soft-voting ensemble | 0.960 (0.900-0.997) | 0.923 | 0.923 | 0.833 | 0.756 |

AUC, area under the receiver operating characteristic curve; MCC, Matthews correlation coefficient. DEG filtering, LASSO/SVM-RFE/Random Forest feature selection, consensus construction, scaling, and model fitting were repeated inside each outer training fold. The 95% CI is a stratified 5000-replicate bootstrap interval for the pooled outer-fold predictions.

**Supplementary Table S4. Clinical and demographic context for the principal RNA-seq and single-cell analyses.**

| **Dataset** | **Group** | **n** | **Age, y mean ± SD** | **BMI mean ± SD** | **HbA1c mean ± SD** | **Female, n** | **Assay** | **Treatment annotation** |
| --- | --- | --- | --- | --- | --- | --- | --- | --- |
| GSE164416 | ND | 18 | 56.7 ± 10.9 | 22.9 ± 2.9 | 5.26 ± 0.31 | not available | bulk RNA-seq | not available |
| GSE164416 | T2D | 39 | 66.9 ± 12.1 | 26.0 ± 5.2 | 7.42 ± 1.29 | not available | bulk RNA-seq | not available |
| GSE221156 beta cells | ND | 17 | 45.4 ± 12.2 | 28.4 ± 5.6 | 5.21 ± 0.29 | 5 | 10x 3' v3: 13; 10x 3' v2: 4 | no antidiabetic treatment expected; field unannotated |
| GSE221156 beta cells | T2D | 17 | 50.4 ± 10.2 | 33.0 ± 5.9 | 7.55 ± 1.72 | 7 | 10x 3' v3: 10; 10x 3' v2: 7 | 7/17 annotated (5 metformin-specific, 2 oral_meds); 10/17 unannotated |

**Supplementary Table S4B. Donor-level clinical metadata for the GSE221156 beta-cell validation cohort. Blank medication entries indicate that no medication annotation was available and must not be interpreted as untreated status.**

| **Donor** | **Group** | **Age** | **BMI** | **HbA1c** | **Sex** | **Assay** | **Medication** | **Beta cells, n** |
| --- | --- | --- | --- | --- | --- | --- | --- | --- |
| HP-20152-01 | ND | 48 | 20.2 | 5.2 | male | 10x 3' v3 |  | 1000 |
| HP18054-01 | ND | 40 | 25.3 | 5.3 | male | 10x 3' v2 |  | 1000 |
| HP18304 | ND | 21 | 27.3 | 5.4 | male | 10x 3' v2 |  | 686 |
| HP19047-01 | ND | 53 | 22.7 | 4.8 | female | 10x 3' v3 |  | 1000 |
| SAMN08768799 | ND | 54 | 35.6 | 5.1 | male | 10x 3' v2 |  | 1000 |
| SAMN09370567 | ND | 32 | 28.5 | 5.2 | male | 10x 3' v2 |  | 1000 |
| SAMN10737781 | ND | 66 | 27.2 | 4.7 | male | 10x 3' v3 |  | 1000 |
| SAMN10873960 | ND | 50 | 24.6 | 5.1 | female | 10x 3' v3 |  | 1000 |
| SAMN11380903 | ND | 37 | 24.2 | 5.5 | male | 10x 3' v3 |  | 1000 |
| SAMN11523048 | ND | 35 | 33.4 | 5.4 | female | 10x 3' v3 |  | 1000 |
| SAMN11633049 | ND | 48 | 38.8 | 5.4 | male | 10x 3' v3 |  | 1000 |
| SAMN11864195 | ND | 53 | 40.0 | 5.5 | female | 10x 3' v3 |  | 1000 |
| SAMN12500521 | ND | 52 | 29.0 | 5.1 | male | 10x 3' v3 |  | 1000 |
| SAMN12673306 | ND | 55 | 28.4 | 5.6 | male | 10x 3' v3 |  | 844 |
| SAMN13108021 | ND | 42 | 25.0 | 5.5 | male | 10x 3' v3 |  | 1000 |
| SAMN15337453 | ND | 26 | 24.2 | 5.2 | male | 10x 3' v3 |  | 1000 |
| SAMN16547514 | ND | 60 | 28.3 | 4.6 | female | 10x 3' v3 |  | 1000 |
| HP17117-01 T2D | T2D | 51 | 35.6 | 7.1 | male | 10x 3' v2 | Oral_meds | 1000 |
| HP1720901T2D | T2D | 52 | 42.8 | 6.6 | female | 10x 3' v2 |  | 1000 |
| HP17225-01T2D | T2D | 59 | 27.7 | 6.5 | male | 10x 3' v2 | Metformin(6yrs) | 833 |
| HP17300-01T2D | T2D | 35 | 27.7 | 6.3 | male | 10x 3' v2 | Metformin(3yrs) | 592 |
| HP18320-01T2D | T2D | 61 | 27.4 | 7.9 | male | 10x 3' v2 |  | 920 |
| HP19044-01T2D | T2D | 37 | 33.0 | 13.1 | female | 10x 3' v3 |  | 1000 |
| SAMN08769026 | T2D | 37 | 38.1 | 8.2 | female | 10x 3' v2 | Oral_meds | 582 |
| SAMN10372788 | T2D | 50 | 35.4 | 9.9 | female | 10x 3' v2 | Metformin | 1000 |
| SAMN10832912 | T2D | 54 | 33.7 | 8.0 | male | 10x 3' v3 |  | 1000 |
| SAMN11157311 | T2D | 34 | 31.7 | 7.3 | female | 10x 3' v3 |  | 1000 |
| SAMN11642375 | T2D | 58 | 32.5 | 6.7 | male | 10x 3' v3 | Metformin(2yrs) | 1000 |
| SAMN12634037 | T2D | 63 | 31.7 | 5.9 | male | 10x 3' v3 | Metformin(5yrs) | 720 |
| SAMN13320559 | T2D | 52 | 24.7 | 6.1 | male | 10x 3' v3 |  | 1000 |
| SAMN13901790 | T2D | 53 | 25.8 | 6.7 | male | 10x 3' v3 |  | 1000 |
| SAMN16365027 | T2D | 62 | 28.5 | 7.6 | female | 10x 3' v3 |  | 1000 |
| SAMN17209843 | T2D | 38 | 43.9 | 7.0 | female | 10x 3' v3 |  | 1000 |
| SAMN17831932 | T2D | 60 | 41.3 | 7.5 | male | 10x 3' v3 |  | 1000 |

Supplementary Table S5. Primary and exploratory donor-level single-cell beta-cell scores in GSE221156. Benjamini-Hochberg q values correct jointly across the seven reported score tests. DiseaseAxis and the exploratory modules are distinct from the primary IsletDysfunctionScore analog.

| **Metric** | **Genes / definition** | **ND mean** | **T2D mean** | **Delta** | **MW p** | **BH q** | **Welch p** | **Interpretation** |
| --- | --- | --- | --- | --- | --- | --- | --- | --- |
| IsletDysfunctionScore analog | z(ImmuneStress) - z(BetaIdentitySecretion) | -0.343 | 0.370 | 0.713 | 0.000944 | 0.00249 | 0.000309 | Primary single-cell analog of the stress-minus-identity framework |
| BetaIdentitySecretion | Expanded 21-gene beta-cell identity/secretion module | 0.8792 | 0.7563 | -0.1229 | 0.000572 | 0.00249 | 0.000207 | Primary identity/secretion validation score |
| ImmuneStress | 14-gene ImmuneStress module | 0.0052 | 0.0074 | 0.0021 | 0.0388 | 0.0543 | 0.0925 | Nominal increase; not significant after BH correction |
| DedifferentiationStress | ALDH1A3, SOX9, HES1, MYC, LDHA, REST, NEUROG3, CDKN1A, CDKN2A | 0.1963 | 0.2669 | 0.0706 | 0.00107 | 0.00249 | 0.000463 | Exploratory dedifferentiation/stress module |
| DiseaseAxis | z(ImmuneStress) + z(ERStress_UPR) + z(DedifferentiationStress) - z(BetaIdentitySecretion) | -0.4425 | 0.4749 | 0.9174 | 0.00979 | 0.0171 | 0.00476 | Exploratory composite; distinct from IsletDysfunctionScore |
| OxidativePhosphorylation | NDUFA1, NDUFB5, NDUFS1, SDHA, SDHB, UQCRC1, COX4I1, COX5A, ATP5F1A, ATP5F1B, ATP5MC1 | 1.1315 | 1.1024 | -0.0291 | 0.0679 | 0.0792 | 0.0556 | Exploratory mitochondrial-energy module |
| ERStress_UPR | HSPA5, XBP1, DDIT3, ATF4, ATF6, EIF2AK3, ERN1, HERPUD1, DNAJB9, PDIA4, EDEM1, SEL1L | 0.7992 | 0.7442 | -0.0550 | 0.0983 | 0.0983 | 0.0631 | Exploratory ER-stress/UPR module |

Medication annotations in GSE221156 were incomplete and did not provide a defined untreated T2D comparator. Accordingly, no causal or drug-specific treatment effect was estimated from these data.

**Supplementary Table S6. Study-level cross-fitted validation of the composite module and expression/detection-matched random gene-set null.**

For each held-out cohort, two 14-gene modules were re-derived using only the other three cohorts. The held-out cohort contributed neither effect direction nor disease labels to module definition. Component and composite Cohen d values are shown. The matched-null p value is the two-sided empirical p for the composite relative to 2000 random 14+14-gene sets matched gene-by-gene for mean expression and nonzero detection rate in the held-out cohort.

| **Held-out cohort** | **Training cohorts** | **n ND** | **n T2D** | **ImmuneStress d** | **Identity d** | **Composite d** | **Matched-null p** |
| --- | --- | --- | --- | --- | --- | --- | --- |
| GSE25724 | GSE20966, GSE38642, GSE164416 | 7 | 6 | 0.503 | -1.076 | 1.012 | 0.3973 |
| GSE20966 | GSE25724, GSE38642, GSE164416 | 10 | 10 | 0.468 | -1.796 | 1.359 | 0.0245 |
| GSE38642 | GSE25724, GSE20966, GSE164416 | 54 | 9 | 0.820 | 0.105 | 0.390 | 0.4188 |
| GSE164416 | GSE25724, GSE20966, GSE38642 | 18 | 39 | 0.828 | -0.192 | 1.129 | 0.0020 |

Random-effects pooling of the four held-out effects: Hedges' g = 0.905 (95% CI 0.501-1.309), p = 1.14 x 10^-5, I2 = 5.5%.

**Supplementary Table S7. Feature-selection stability across the five outer folds of fully nested cross-validation.**

Frequencies are reported for the 10 features in the final full-cohort candidate panel; selection in each outer fold was performed using only that fold's training data.

| **Final-panel feature** | **Outer folds selected (of 5)** | **Selection frequency** |
| --- | --- | --- |
| GABRA2 | 5 | 1.0 |
| ENSG00000284653 | 5 | 1.0 |
| PRIMA1 | 3 | 0.6 |
| RNU1-70P | 2 | 0.4 |
| SLC2A2 | 4 | 0.8 |
| ARG2 | 3 | 0.6 |
| DKK3 | 2 | 0.4 |
| TAFA4 | 1 | 0.2 |
| HHATL | 3 | 0.6 |
| ENSG00000227589 | 3 | 0.6 |
